# Limits of Trial-Adaptive Neural–Language Fusion Across Large Language Models in P300 Brain–Computer Interfaces

**DOI:** 10.64898/2026.08.30.26361777

**Authors:** Alon Gorenshtein, Mahmud Omar, Eric Jia, Yosef Adiniaev, Oved Daniel, Jonathan Kruskal, Muneeb Ahmed, Olga Brook, Eyal Klang, Yiftach Barash

## Abstract

**Objective:** Published P300-speller fusion schemes fix prior trust regardless of trial reliability; we tested whether a reliability estimate improves on it.

**Methods:** We reanalyzed 3,373 archived P300-speller selections from 47 people with ALS (BigP3BCI). A fair, matched-search-space comparison, tuning both a fixed weight and an adaptive policy out-of-fold, was evaluated across 22 evaluable language-model priors up to 46.7B parameters. Two representative priors, GPT-2 and a classical 5-gram, additionally received detailed naive and mechanistic analyses.

**Results:** No prior’s 95% CI favored adaptive fusion under the fair comparison, despite unexploited oracle headroom at every scale. Under GPT-2, the naive comparison was significantly worse for adaptive fusion; both anchors converged to a degenerate or near-degenerate fair-comparison solution. For the representative anchors, three further controllers failed to convert that headroom into benefit; the fixed-fused posterior’s output probability outperformed the best controller for flagging errors (2.8- to 3.8-fold enrichment).

**Conclusion:** A tuned fixed weight is a difficult-to-beat default across the tested scale range; reliability estimation gave no deployable adaptive advantage.

**Significance:** Adaptive weighting should be validated against a fairly tuned baseline across model families and scales; in this dataset, the fused output’s confidence identified high-risk selections better than the tested purpose-built ranker.

## I. Introduction

**P**300 spellers let people with severe motor impairment, including amyotrophic lateral sclerosis (ALS), communicate by decoding an event-related brain response to a target symbol [9]. Because written language is structured, several groups have improved speller accuracy by fusing the neural posterior with a language-model prior: classical n-gram and word-level priors combined with the neural likelihood through a fixed Bayesian product or a particle-filter structure [4], [5], [6], and, more recently, generative large language models substituted for the classical prior [10], [11]. Separately, dynamic-stopping methods already estimate how reliable the accumulating neural evidence is on a given trial, using that estimate to decide when to stop collecting flashes rather than how much to trust the language model [5], [12].

Published fusion schemes, classical or generative, generally set the language model’s influence at a fixed weight or a fixed Bayes-rule product, independent of how reliable the neural evidence is on that particular trial [4], [5], [6], [7], [10], [11], [13]. A 2025 perspective on integrating large language models into brain–computer interfaces names the integration of neural decoding outputs with probabilistic language generation an unresolved challenge rather than a solved one [13]. A related approach in continuous brain-to-text decoding calibrates neural-decoder uncertainty to drive language-model integration and error-flagging, but for phoneme-level speech decoding through hypothesis-level control, not for discrete character-by-character selection through a continuously tunable per-trial weight [14]. Two recent online P300-speller systems integrate a large language model directly into the interface, one using it to drive word-level sentence-composition prediction and the other adding a locally deployed, ALS-adapted model with an adaptive stimulus-acquisition strategy, and both report substantial gains in communication rate [15], [16]. Neither, however, continuously adapts the relative weight given to the neural posterior versus the language-model prior on a per-selection basis, the specific mechanism this study tests.

Whether a trial-level estimate of neural reliability, derived only from the shape of the neural posterior itself and validated against realized outcomes, can set a language model’s fusion weight better than a fixed weight has not been tested. We conducted a retrospective reanalysis of an existing online P300-speller archive, comparing a reliability-gated adaptive fusion policy against a no-language-model floor, fixed-weight fusion, and a grid oracle upper bound informed by each trial’s own true outcome, on the net correctness the language-model prior added relative to the neural posterior alone, across 22 evaluable language-model priors spanning classical n-grams to 46.7-billion-parameter causal language models. Two representative priors additionally received deep mechanistic characterization.

## II. Methods

### A. Study Design, Data, and Ethics

This was a retrospective secondary reanalysis of neural posteriors and language-model prior posteriors already computed by a companion, same-laboratory attribution study on the same archive (unpublished). No new EEG decoding was performed anywhere in this study; the decoded-history analysis (Section III-G) instead required additional forward passes of the existing language models on newly decoded contexts. Data came from BigP3BCI v1.0.0, a public, CC-BY-4.0 archive of legacy online P300-speller studies in people with ALS [3]; the original studies were approved by the relevant institutional review boards, with informed consent obtained from all participants or their legally authorized representatives. Source studies B, F, L, and N were included: 3,373 online selections from 47 participants across 115 sessions, spanning 951 unique linguistic contexts, each with a 36-way neural posterior and, for each of 24 non-null language-model priors, a matching 36-way prior posterior conditioned on the ground-truth copy-spelling prefix. This reanalysis used only previously collected, de-identified, publicly available records and required no new institutional review board approval or informed consent. Reporting followed STROBE [1] and an LLM-BCI reporting checklist [2].

### B. Neural Posteriors, Language-Model Priors, and Fusion

Neural posteriors were the companion study’s session-specific logistic decoders, fit from each session’s calibration phase only. Priors spanned three classical character 5-grams and 21 causal language models across ten families (124M–46.7B parameters), each scored once per context by marginalizing its next-token distribution onto the 36-symbol grid alphabet. Of 24 non-null priors, 2 (gpt2-large, google/recurrentgemma-2b) could not be evaluated under the fair, matched-search-space procedure (Limitations), leaving 22 evaluable priors – 19 causal language models spanning 124M–46.7B parameters and 3 classical n-grams – for the principal exploratory cross-model analysis, using the identical procedure for every prior. GPT-2 (general-purpose) and a Kneser–Ney-smoothed 5-gram (classical baseline, hereafter 5-gram) were carried through every analysis as representative anchors. All 24 non-null priors, including the 2 unevaluable by the fair comparison, entered a naive ladder-wide consistency sweep; 3 representative priors additionally entered a candidate-arbitration extension. Both sources were combined by the standard, uncalibrated Bayesian speller product: the fused posterior is proportional to the neural posterior raised to a fusion exponent *β*, times the prior posterior; *β* = 1 is the field default [4], [5], [6], [7].

### C. Trial-Level Reliability Estimator and Adaptive Fusion

For each selection, three features were computed from the neural posterior alone (no access to the true target): Shannon entropy, top-1 probability, and top-1-to-top-2 margin. An L2-regularized logistic regression mapped these to *ĝ*, an out-of-fold estimate (5-fold, grouped by participant) of the probability the neural argmax is correct. *ĝ* set a fusion weight via *β* = *β*_min_ *·* (*β*_max_*/β*_min_)^*ĝ*^, with *β*_min_ = 0.25 and *β*_max_ = 4.0, the endpoints of the existing fixed-*β* grid.

### D. Comparison Arms

Four primary arms were compared: a no-language-model floor (neural argmax only), fixed-weight fusion (*β* = 1), adaptive fusion (*β* set per selection from *ĝ*), and a grid oracle upper bound (*β* chosen from {0.25, 0.5, 1, 2, 4} using each trial’s own already-known outcome). Because *β* = 1 was never claimed the best achievable fixed value, a second, fairer comparison tuned both sides out-of-fold: a best-fixed-*β* control selected the single best grid value per fold from the other folds’ participants, and a tuned-adaptive arm selected a (*β*_min_, *β*_max_) anchor from a 12-anchor grid that fully nests every value the fixed control could choose (a degenerate anchor *β*_min_ = *β*_max_ = *b* for every *b* in the fixed grid, plus 7 non-degenerate ranges), so the adaptive arm’s search space is a strict superset of the control’s, never a subset.

### E. Outcomes and Statistics

Prior capture was initially specified as the principal end-point; after its one-sided optimum was identified (favoring disabling the language model regardless of merit; Discussion, Limitations), net correct gain was designated the principal exploratory endpoint instead. Net correct gain is the fused posterior’s correctness minus the neural posterior’s own correctness (−1, 0, +1) per selection; its mean equals phantom agreement minus prior capture (Supplementary Material). Arm contrasts on net correct gain used a participant-cluster bootstrap (2,000 resamples, 95% percentile CI); the three binary secondary outcomes used participant-clustered GEE logistic models (odds ratios, Benjamini–Hochberg-adjusted *P* values [8]). Transportability (leave-one-study-out) and a 24-prior naive ladder-wide sweep assessed generalizability (Section III). The fair, matched-search-space comparison, the principal cross-model analysis, covered all 22 evaluable priors using the identical procedure throughout (Section III-B); the candidate-arbitration controller was separately extended to 3 further representative priors using the identical procedures above; gpt2-large and google/recurrentgemma-2b could not be evaluated by the fair comparison (Section IV, Limitations).

### F. Three Further Per-Trial Controllers

Three subsequent, distinct designs each tried to convert *ĝ*’s predictive signal (Section III-D) into deployable benefit, each evaluated on its own held-out data with full detail in the Supplementary Material. (i) A joint neural-and-language-model gate combined both posteriors’ raw features into a fusion-weight override, held out on one source study never touched during development. (ii) An actionability-aware controller, motivated by most of that gate’s deviations being outcome-irrelevant, restricted overrides to selections its own classifier judged decision-sensitive, held out on a second study. (iii) A candidate-arbitration controller worked directly in each selection’s small, grid-derived set of reachable candidates (the fused argmax at each grid *β*, unioned with the neural-only argmax), using boundary-distance features exact within that candidate set to decide whether to override the fixed winner; it was evaluated by nested leave-one-study-out cross-validation across all four studies. A final check repurposed the candidate ranker’s own confidence score, and seven simpler baseline signals, for selective error flagging rather than overriding (Section III-F; Supplementary Material S1.11).

## III. Results

### A. Cohort

The analysis included 3,373 online P300-speller selections from 47 participants with ALS across 115 sessions and four source studies. The neural posterior’s own argmax was correct in 71.8% of selections, identical across arms and priors since it is independent of the language model. Of 24 non-null priors, 2 (gpt2-large, google/recurrentgemma-2b) could not be evaluated under the fair, matched-search-space procedure, leaving 22 evaluable priors as the principal exploratory cross-model analysis; GPT-2 and a Kneser–Ney-smoothed 5-gram served as representative anchors for full mechanistic characterization; all 24 priors entered a simpler naive ladder-wide sweep (Section II-B).

### B. No Prior Favored Adaptive Fusion Across the Model Ladder

A naive (*β* = 1) sweep across all 24 non-null priors found no prior favoring adaptive fusion on prior capture (5 of 24 favored fixed; the rest included zero) and a more pronounced fixed-favoring pattern on fused selection correctness. The fair, matched-search-space comparison, the principal cross-model analysis, covered all 22 evaluable priors (Section II-B): 19 causal language models and 3 n-grams, 124M–46.7B parameters. No prior’s 95% CI excluded zero favoring adaptive fusion on net correct gain (9/22 exact ties, 2/22 excluded zero favoring fixed, 11/22 crossed zero), still uncorrected for multiplicity, and the grid oracle exceeded the best-tuned fixed weight by 2.7–6.7 percentage points at every prior (eTable15; Fig. 1); the two anchors are among these 22, detailed next (Section III-C). The candidate-arbitration controller, extended to three further priors (1.5B, 12B, 46.7B parameters), showed the same net loss as the original two (−0.0033 to −0.0080; eTable12).

**Fig. 1.**
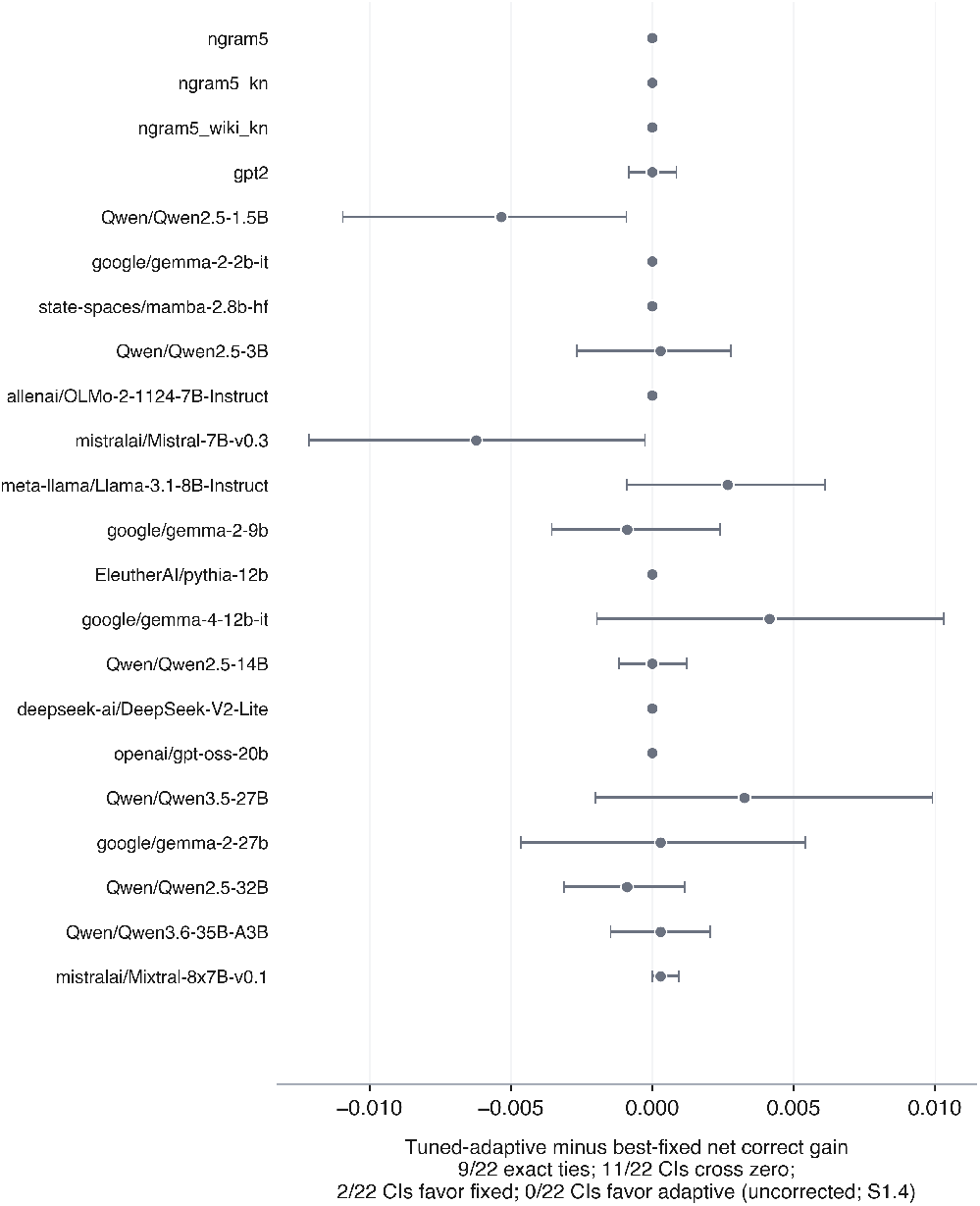
No prior favored adaptive fusion across the full model ladder. The fair, matched-search-space comparison across all 22 evaluable priors, sorted by parameter count ascending, from a classical n-gram to a 46.7-billion-parameter causal language model. Tuned-adaptive-minus-best-fixed difference in net correct gain for each prior; error bars are 95% participant-cluster bootstrap confidence intervals, uncorrected for multiplicity across these 22 comparisons. All points are drawn in one neutral color; the panel reports the exact-tie, CIs-cross-zero, favors-fixed, and favors-adaptive counts directly, since these comparisons are read descriptively, not as independent significance tests.

### C. The Two Representative Anchors

Adaptive fusion did not exceed fixed-weight fusion on net correct gain for either anchor (Fig. 2a–b). Under GPT-2, mean net correct gain was 0.0193 for fixed and 0.0125 for adaptive, a difference of 0.0068 (95% CI, −0.0128 to −0.0012), favoring fixed weighting; under 5-gram the gap was smaller and non-significant (−0.0039; 95% CI, −0.0096 to −0.0021). Under the fair, matched-search-space comparison, in which the adaptive search space fully contained the fixed control’s own options, the tuned-adaptive arm still did not beat the best-fixed-*β* control on any outcome, converging to a degenerate or near-degenerate solution instead (Fig. 2c–d). On prior capture, both arms selected the fully degenerate *β* = 4 in every fold for both priors. On fused selection correctness and net correct gain the tie was exact at the point estimate for both priors: a true selection-by-selection identity for 5-gram, every fold choosing a degenerate anchor matching the control exactly, but a near-degenerate (0.9, 1.1) anchor in some GPT-2 folds, which is why only its interval has nonzero width. The grid oracle, restricted to the same five-value grid, exceeded fixed-weight fusion by 0.0519 (95% CI, 0.0382–0.0655) under GPT-2 and 0.0421 (95% CI, 0.0286–0.0563) under 5-gram, showing headroom that neither naive nor fairly-tuned adaptive weighting reached.

**Fig. 2.**
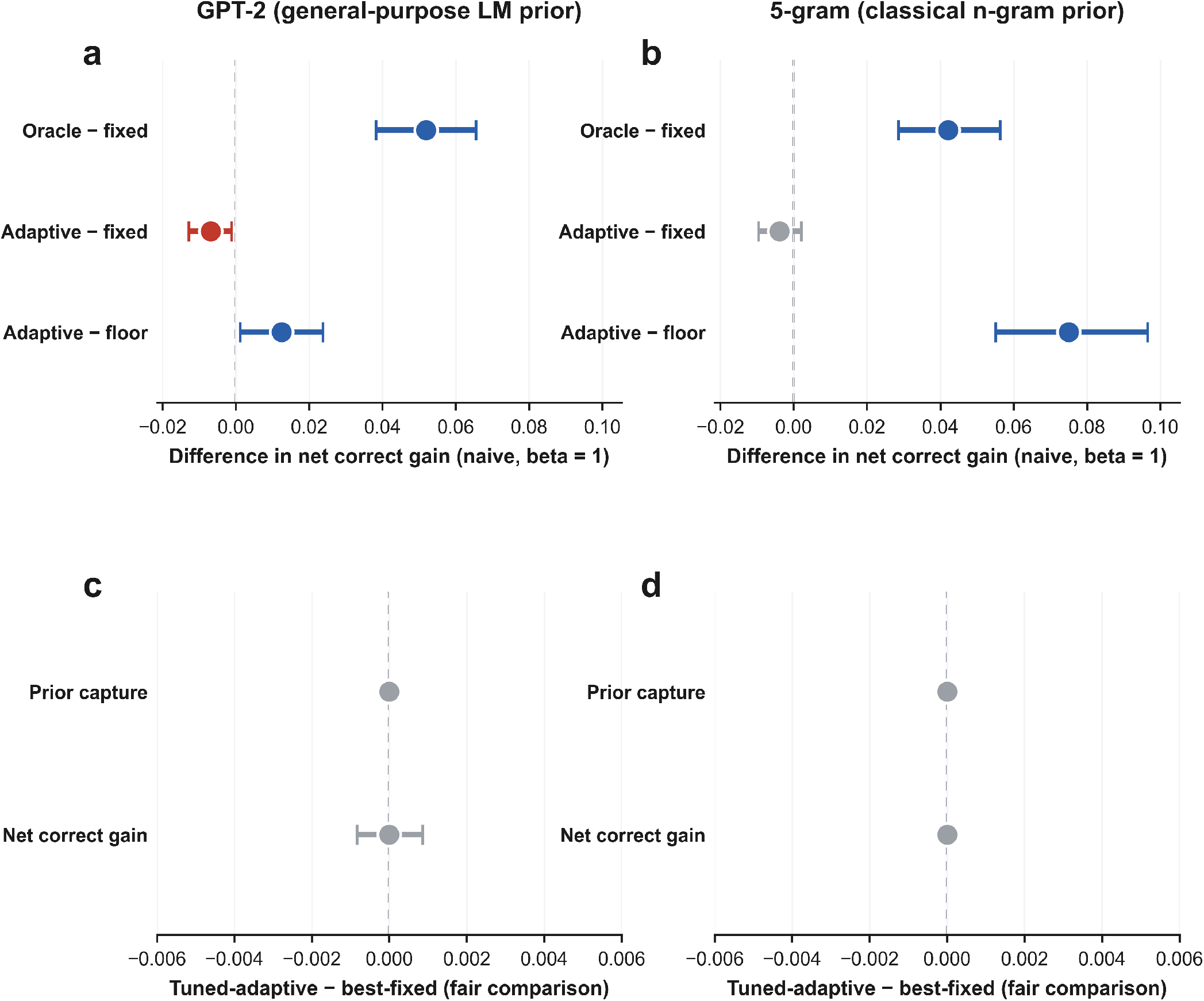
Detailed characterization of the two representative anchors did not favor adaptive fusion. (a,b) The naive comparison against the field-default fixed weight (*β* = 1), for GPT-2 (a) and the 5-gram (b). (c,d) The fair, matched-search-space comparison, in which both the fixed control and the adaptive policy were tuned out-of-fold, the adaptive search space fully containing every option available to the fixed control, for GPT-2 (c) and the 5-gram (d); the adaptive search converged to a degenerate or near-degenerate solution that did not improve on best-fixed fusion for either prior.

### D. The Reliability Estimate Predicts Correctness but Has Little to Act On

The reliability estimator discriminated correct from incorrect neural selections with an out-of-fold AUROC of 0.845 and expected calibration error of 0.050 (eFigure 2), identical across priors since it is fit from the neural posterior alone.

That signal, however, has little decision space to act on: an audit of each selection’s grid-achievable decoded candidates found that most admit only one (66.85% GPT-2, 78.42% 5-gram; Fig. 3a), leaving a well-calibrated per-trial signal with no lever to pull in most selections. This grid-level constraint, not the estimator’s failure, may help explain why fair tuning favored degenerate or near-degenerate adaptive solutions rather than exceeding fixed-weight fusion (Section III-C).

**Fig. 3.**
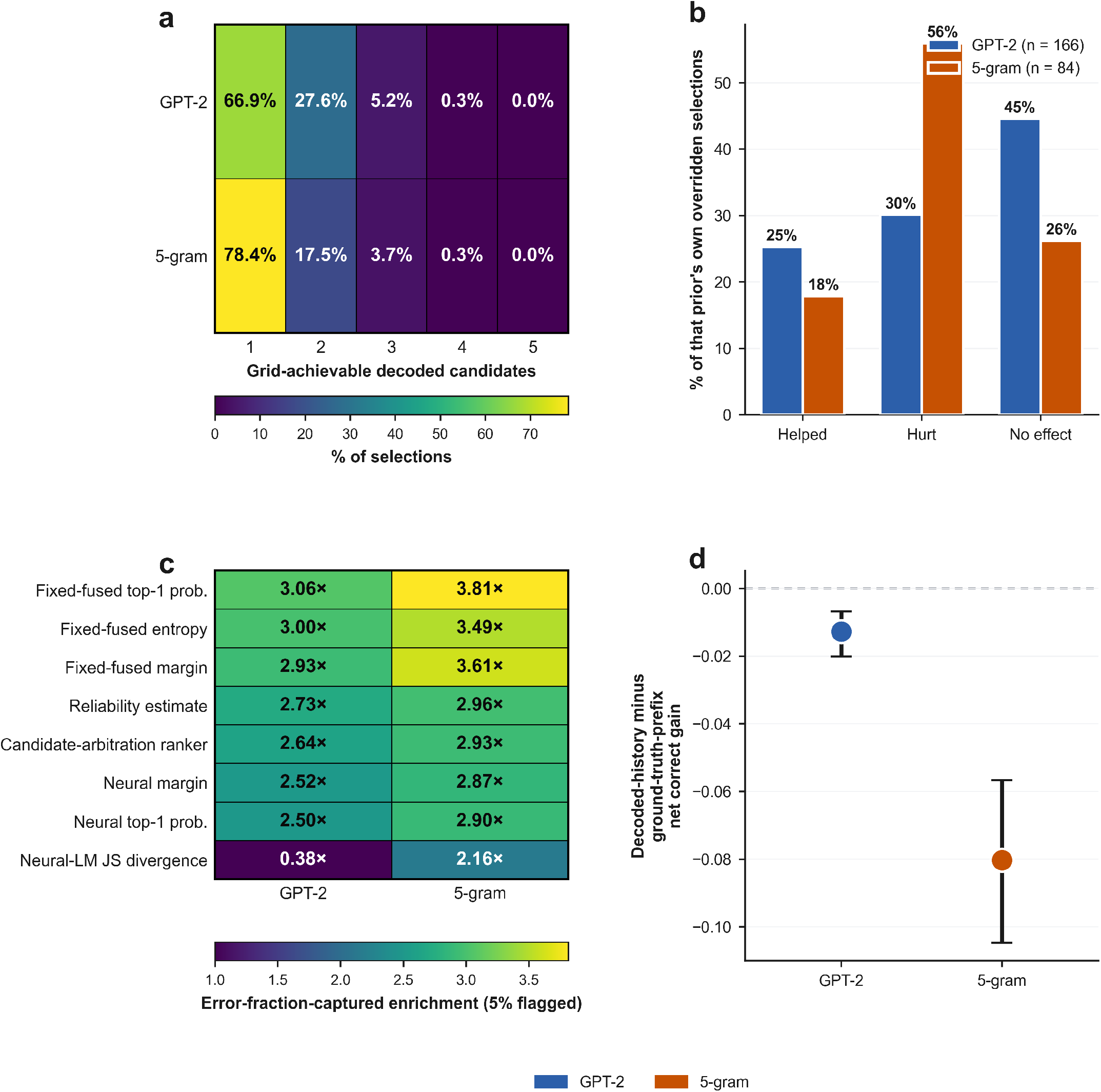
Mechanistic basis for the collapse and the deployment implications. (a) Fraction of selections with 1–5 grid-achievable decoded candidates across neural-only decoding and the fixed-weight fusion grid, both priors. (b) Candidate-arbitration override outcomes among overridden selections, as a percentage of that prior’s own overridden total (GPT-2, *n* = 166; 5-gram, *n* = 84). (c) Error-fraction-captured enrichment at 5% flagged for all eight benchmarked confidence signals, both priors. (d) Decoded-history-minus-ground-truth-prefix difference in net correct gain, both priors, with 95% participant-cluster bootstrap confidence intervals.

### E. Three Further Controllers Did Not Convert That Signal Into Benefit

Three subsequent, distinct per-trial designs each tried to convert the reliability signal into deployable benefit for the minority of selections where it could matter; none did. A joint neural-and-language-model gate predicted the grid oracle’s own chosen weight with real discriminative signal (macro AUROC, 0.79 GPT-2, 0.81 5-gram) but, evaluated on a held-out study, did not exceed best-fixed-weight fusion (net correct gain difference: GPT-2, −0.0128, 95% CI, −0.0271 to 0.0000; 5-gram, −0.0106, 95% CI, −0.0212 to −0.0021). An actionability-aware controller, restricted to selections judged decision-sensitive, also did not beat best-fixed-weight fusion on its own held-out study (GPT-2, −0.0035, 95% CI, −0.0071 to 0.0000; 5-gram, +0.0012, 95% CI, −0.0033 to 0.0050). A candidate-arbitration controller, working in each selection’s own small, grid-derived candidate set with boundary-distance features exact within that set, was evaluated by nested leave-one-study-out cross-validation across all four studies: the difference in net correct gain versus best-fixed fusion was −0.00237 for GPT-2 (95% CI, −0.0085 to 0.0031) and −0.00949 for 5-gram (95% CI, −0.0148 to −0.0042), a net loss under both priors, significant for 5-gram. Among overridden selections, more were hurt than helped for both priors (Fig. 3b).

### F. Flagging by a Simpler Signal Is More Tractable

The candidate-arbitration controller’s own confidence prompted a check for selective error flagging. Benchmarked against seven alternative signals, the fixed-fused posterior’s own top-1 probability ranked first or second at both bench-marked coverage levels and both priors (Fig. 3c). At 5% flagged, it captured a disproportionate share of the fixed comparator’s own errors (GPT-2, 3.06-fold enrichment; 5-gram, 3.81-fold); at 10% flagged, GPT-2, 2.78-fold and 5-gram, 3.39-fold. The candidate ranker itself was not better at either coverage level and was lower on enrichment in the paired-bootstrap comparison for both priors, with the 95% CIs excluding zero: the flagging capability traces to the existing fused output, not to added ranker complexity.

### G. Secondary Checks

Leave-one-study-out transportability favored fixed-weight fusion in most folds (prior capture: 5 of 8; fused selection correctness similar), significant in only 1 of 8 for each outcome. Because every analysis above conditioned each prior on the ground-truth copy-spelling prefix, a final check replayed each prior at its own frozen best-fixed weight on the fusion arm’s own previously decoded symbols instead; this materially worsened net correct gain for both priors (GPT-2, −0.0127, 95% CI, −0.0201 to −0.0068; 5-gram, −0.0803, 95% CI, −0.1048 to −0.0566), roughly six times larger in absolute terms for the classical prior (Fig. 3d), showing ground-truth-prefix evaluation can overestimate deployable fusion benefit. Full secondary-outcome statistics (prior capture, phantom agreement, overall fused selection correctness) are in the Supplementary Material.

## IV. Discussion

In this retrospective reanalysis of an online P300-speller archive, a trial-level neural-reliability estimate did not improve language-model fusion beyond a single fixed weight, across a fair, matched-search-space comparison spanning 22 evaluable priors: no prior’s CI favored adaptive fusion, with unexploited oracle headroom at every scale tested. For two representative anchors, GPT-2 and a classical 5-gram, the same fair, out-of-fold-tuned comparison, which fully contained the fixed control’s own options, converged to a degenerate or near-degenerate solution rather than exceeding it, for both outcomes. Under a naive, un-tuned comparison, adaptive fusion was measurably worse than fixed-weight fusion under GPT-2.

The reliability estimator carries real information about neural trustworthiness (out-of-fold AUROC, 0.845), yet most selections leave that information nowhere to act: most admit only one grid-achievable decoded candidate, so no grid value of the weight changes the decoded symbol. The grid oracle, using each trial’s own true outcome and undeployable, still exceeded fixed-weight fusion by a material margin, showing real headroom concentrated in the minority of selections with more than one grid-achievable candidate.

Three subsequent, distinct per-trial designs each tried to convert that headroom into deployable benefit, and none did. Oracle-predictive signal (from a joint neural-and-language-model gate) did not translate into a deployable policy on held-out data. Restricting a controller to only the decision-sensitive minority did not exceed best-fixed-weight fusion on its own held-out study. A candidate-arbitration controller, this project’s most targeted per-trial mechanism yet, again did not beat best-fixed-weight fusion under nested leave-one-study-out cross-validation, significantly worse for 5-gram. A simpler, different signal was useful instead: the fixed-fused posterior’s own top-1 probability, requiring no additional fitting, outperformed the candidate-arbitration ranker for flagging the fixed comparator’s own errors – the flagging capability traces to the existing fused output, not to added controller complexity.

Prior fusion schemes fix the language model’s weight independent of per-trial neural reliability, from classical n-gram and Bayesian speller fusion to generative large-language-model systems [4], [5], [6], [7], [10], [11]. Dynamic-stopping methods estimate online neural reliability to decide how many flashes to collect, not how much to trust a language-model prior [5], [12]. A recent brain-to-text preprint uses decoder uncertainty for phoneme-level speech decoding through hypothesis-level re-ranking [14]; this study instead tests uncertainty as a continuously tunable weight for discrete, per-character P300 selection, and finds a simple, entropy-and-margin-based version does not improve fusion.

For groups building or deploying language-model fusion in P300 spellers, these findings support a fixed, out-of-fold-tuned weight as a simple, difficult-to-beat default over posterior-shape-based adaptive weighting on this evidence. Where flagging low-confidence selections for review, rather than reweighting, is desired, the fixed-fused posterior’s own output probability is the better-supported signal.

### A. Limitations

This was a retrospective reanalysis with no prospective closed-loop deployment; the reliability estimator used only three posterior-shape features (entropy, top-1 probability, margin), so richer signals (EEG amplitude, session recency, fatigue) were not tested; the cohort is a single archive of four ALS studies on one montage and paradigm; prior capture, the originally specified principal endpoint, is one-sided (rewards only harmful overrides) and was replaced by net correct gain after this was discovered, a disclosed deviation, so no part of this study is confirmatory in the preregistered sense; cross-validation and inference clustered only on participant, and the 3,373 selections span only 951 unique linguistic contexts; the 24-prior sweep, the 22-prior fair comparison, and error-flagging comparisons were uncorrected for multiplicity; gpt2-large’s posterior did not match the companion-study scoring run and google/recurrentgemma-2b was absent from the available scored data, excluding both from the fair comparison, so the principal exploratory cross-model analysis spans 22, not 24, priors, though both entered the naive ladder-wide sweep – this 22-prior analysis was itself added after the two-anchor results, a further disclosed deviation from a preregistered design; each per-trial controller’s holdout is drawn from the same archive and paradigm, not an external site, and by the candidate-arbitration stage every source study had already been touched by an earlier controller’s development, so its leave-one-study-out evaluation is exploratory, not independent or prospective; the candidate audit and candidate-arbitration controller both define candidates from the fixed-weight grid {0.25, 0.5, 1, 2, 4} plus the neural-only argmax, so the true count of continuously reachable candidates between grid points was not exactly enumerated and could exceed the grid-achievable count reported here; and the derived posterior matrices behind this analysis are not yet publicly redistributed, though we commit to a Zenodo deposit with a citable DOI upon acceptance (Data and Code Availability). Methodological detail behind each limitation is in the Supplementary Material (S1.6–S1.11).

## V. Conclusion

A single, well-tuned fixed fusion weight was not beaten by a trial-level, posterior-shape-based reliability-gated adaptive policy in this reanalysis, across a fair, matched-search-space comparison spanning 22 evaluable priors, a transportability check, a 24-prior naive sweep, and, for two representative anchors, three subsequent, distinct per-trial control designs. Real headroom remains in the minority of selections with more than one grid-achievable candidate, but none of three designs converted it into net benefit; a simpler confidence signal, the fixed system’s own output probability, was useful for identifying error-enriched selections instead. These results motivate a search for what separates predictive signal from a deployable arbitration policy, and support flagging low-confidence selections for review as a more conservative use of that signal than acting on it directly, rather than abandoning per-trial fusion adaptation as a concept.

## Data Availability

All data produced are available online at github.com/Alon-Gorenshtein/LLM-BCI-Fusion

## Data and Code Availability

Analysis code, exact seeds, and figure/table scripts are at github.com/Alon-Gorenshtein/LLM-BCI-Fusion. Selections and posteriors are read-only inputs from the companion attribution study; they will be deposited in a public Zenodo archive with a citable DOI upon acceptance. BigP3BCI access requires no data use agreement.

## Acknowledgment

The authors declare no competing interests. Conception and study design: A.G. Data curation and analysis: A.G. Drafting the manuscript: A.G. Critical revision for important intellectual content: all authors. Supervision: E.K., Y.B.

