## appendix for "Limits of Trial-Adaptive Neural–Language Fusion Across Large Language Models in P300 Brain–Computer Interfaces"

This Supplementary Information accompanies the main manuscript. It contains the full reliability- estimator and fusion-arm implementation detail, the fair-comparison (rescue) search-space design, the per-arm and per-prior result tables, and the transportability and consistency-sweep figures summarized in the main text.

**Contents**

- S1 Supplementary Methods (S1.1 to S1.11)
- S2 Supplementary Tables (eTable 1 to eTable 14, plus eTable 4b and 5b)
- S3 Supplementary Figures (eFigure 1 to eFigure 4)
- S4 Code and Data Availability

### S1 Supplementary Methods

#### S1.1 Reliability features

Three features were computed from each selection's 36-way neural posterior, without reference to the true target symbol: Shannon entropy in bits, the top-1 probability, and the top-1-to-top-2 margin (the top-ranked symbol's probability minus the runner-up's). These three features, and only these three, were the input to the reliability estimator; no raw EEG features, session covariates, or timing information entered the estimator.

#### S1.2 Reliability estimator and the adaptive link

An L2-regularized logistic regression (scikit-learn defaults, random state 20260820) mapped the three reliability features to ĝ, an estimated probability that the neural posterior's own argmax was correct. Fitting used 5-fold grouped cross-validation (participant as the group), so every ĝ reported anywhere in this study is out-of-fold: no participant's own rows ever contributed to the model that scored that participant's selections. ĝ was mapped to a fusion weight through beta = beta_min times (beta_max / beta_min) raised to the power ĝ, with beta_min = 0.25 and beta_max = 4.0, the endpoints of the existing fixed-beta grid {0.25, 0.5, 1, 2, 4} used throughout this study and the companion attribution study, so the adaptive arm's weight range never exceeds the range already validated for fixed-weight fusion.

#### S1.3 The rescue (fair-comparison) search space

The original four-arm comparison (no-language-model floor, fixed-weight fusion at beta = 1, adaptive fusion, oracle) found the adaptive arm losing to fixed-weight fusion. Because beta = 1 was never claimed to be the best achievable fixed weight, a second, more stringent comparison let both a fixed-beta control and the adaptive policy's own tuning parameters be chosen out-of-fold, so that neither side had an unfair advantage.

The fixed-beta control (best_fixed) chose, for each of 5 participant-grouped folds, whichever grid beta in {0.25, 0.5, 1, 2, 4} performed best on the outcome under test using only the other folds' participants, then applied that beta's precomputed row to the held-out fold.

The tuned-adaptive arm fit a fresh reliability estimator on each fold's training participants only (not a single estimator shared across folds), then chose a (beta_min, beta_max) anchor pair for the adaptive link from a grid of 12 candidates: one degenerate anchor (beta_min = beta_max = b) for every b in {0.25, 0.5, 1, 2, 4}, so every fixed-beta control value is exactly reachable by the adaptive arm, plus 7 additional non-degenerate anchors ((0.9, 1.1), (0.8, 1.25), (0.5, 2.0), (0.25, 4.0), (0.1, 4.0), (0.1, 20.0), (0.5, 8.0)) spanning narrower and wider ranges around beta = 1. The anchor was chosen using the same fold's training participants' in-sample ĝ predictions, then applied to the held-out fold. This anchor-selection step uses in-sample predictions from that fold's own freshly fit estimator, not a further nested inner-cross-validation split; a direct comparison on this dataset found the in-sample and a true nested-inner-CV anchor choice agreed almost perfectly (correlation, 0.998), so this simplification was retained, described accurately rather than represented as fully nested cross-validation. No test participant's own data ever informed its own fold's estimator fit or its own fold's anchor choice.

The anchor grid must include a degenerate option matching every value the fixed-beta control could choose, or the comparison would be biased against the adaptive arm whenever the control selected a value the adaptive side could not reach; the control did choose different grid values across outcomes and priors (beta = 4 for prior capture under both priors; beta = 0.25 and 0.5 for fused correctness under ngram5_kn), so this fully nesting property was necessary, not incidental, to a fair comparison.

#### S1.4 Ladder-wide consistency sweep

The 24-prior sweep (eTable 4) used the same non-null priors as the companion attribution study's own prior ladder, read directly from that study's attribution table (prior_model column) rather than from the prior-shard file names, because several model identifiers contain a forward slash that the file-naming convention escapes to an underscore, and re-deriving the canonical name from the escaped file name is not always reversible. This sweep is a consistency check across the ladder rather than a confirmatory hypothesis-test family; no correction for the 24 comparisons was applied to eTable4 or eTable4b, and the per-prior intervals in those tables should be read descriptively rather than as an independent significance test for each prior.

The fair, matched-search-space comparison (rescue analysis; S1.7) was similarly extended to every non-null prior except gpt2-large and google/recurrentgemma-2b, which could not be evaluated (Limitations), using the identical nested-cross-validation procedure applied throughout, including a re-validation of the published ngram5_kn result, which reproduced exactly. eTable15 and Figure 1 (main text) report all 22 evaluable priors as one unique set -- the 20 priors newly added by this extension plus gpt2 and ngram5_kn's own already-published rescue results (S1.7), not shown as a separate 21-prior extension; no correction for the 22 simultaneous comparisons was applied, matching eTable4/eTable4b's own convention above.

#### S1.5 Software and reproducibility

All analyses used Python 3.9 with pandas, NumPy, scikit-learn, and statsmodels. All cross-validation splits and bootstrap resamples used fixed random seeds (20260820 for the reliability estimator, the rescue analysis, and the transportability analysis; the same value seeds the participant-cluster bootstrap in every reported contrast). The generalized estimating equations for the three binary secondary outcomes used an independence working correlation (the statsmodels default), not an exchangeable structure; this is stated explicitly because the companion attribution study's own statistical convention uses an exchangeable structure, and the two should not be assumed identical when comparing methods across the two papers.

#### S1.6 Joint neural-and-language-model gate (supplementary analysis)

Study N (8 participants, 470 selections) was reserved as a holdout before any part of this supplementary analysis began. Every step below except the final evaluation used only Studies B, F, and L; Study N was read exactly once, for the final evaluation, and never for calibration, feature design, the oracle-structure check, or gate training.

**Calibration.** A scalar temperature was fit per source (neural, language model) by grid search (25 points, geometrically spaced from 0.2 to 5.0) over 5-fold participant-grouped cross-validation, minimizing mean negative log-likelihood of the true target symbol on the held-out fold; the calibrated posterior is proportional to the raw posterior raised to the power 1/temperature, renormalized. Fitted temperatures: neural, 3.18 (identical across priors, since the neural posterior does not depend on the language-model prior); language model, 2.24 under gpt2 and 1.18 under ngram5_kn. All three exceed 1, indicating every source's raw posterior was overconfident relative to its own accuracy. The existing four-arm comparison was repeated with both posteriors calibrated before fusion; the adaptive arm's own reliability estimator was refit on the calibrated neural posterior (eTable 8).

**Joint features.** Fifteen features were computed from each selection's pair of raw (uncalibrated) 36-way posteriors, without reference to the true target: each source's Shannon entropy, top-1 probability, and top-1-to-top-2 margin; whether the two sources' argmax symbols agreed; each source's probability mass on the other source's argmax; each source's rank of the other source's argmax (0 if it is that source's own argmax); the Kullback-Leibler divergence of the neural posterior from the language-model posterior; the Jensen-Shannon divergence between the two; and the difference and ratio of the two sources' entropies.

**Oracle-structure check.** Before building a gate, a participant-grouped, out-of-fold multinomial logistic regression was fit to predict the grid oracle's chosen beta (S1.3) from the fifteen joint features, on Studies B, F, and L only (2,903 selections). Macro one-vs-rest area under the curve was 0.79 for gpt2 and 0.81 for ngram5_kn, both well above a pre-specified threshold of 0.55, so the analysis proceeded to building a gate.

**Gate.** Two logistic regressions (each with per-fold feature standardization fit on the training fold only) were fit by 5-fold participant-grouped cross-validation on Studies B, F, and L to predict, out-of-fold, phantom agreement and prior capture from the fifteen joint features at the field-default weight (beta = 1); their difference is V(x), an estimate of the net benefit of language-model influence for that selection. The fusion weight defaulted to the best fixed weight (chosen, like the rescue analysis's fixed-beta control, from {0.25, 0.5, 1, 2, 4} using all of Studies B, F, and L) and deviated to 0.5 or 2 times that weight when V(x) crossed a threshold (high, low), a pair searched over {0.02, 0.05, 0.1, 0.2} for high and its negatives for low, selecting the pair that maximized gated fused selection correctness on Studies B, F, and L. This grid's largest magnitude is 0.2, so no (high, low) pair reproduces exact identity to the best fixed weight; the widest, most-deferring pair, (0.2, -0.2), was available in the search and was not selected for either prior, which instead chose a narrower band on one side (gpt2: high = 0.02, low = -0.2; ngram5_kn: high = 0.2, low = -0.02).

**Frozen evaluation.** The selected (best fixed weight, high, low) triple is the single frozen policy. Its net-benefit model was refit once on all of Studies B, F, and L (no cross-validation, since Study N is not itself split further) and applied to Study N's raw joint features only; Study N's own outcome labels were never read by this refit. The frozen policy did not clearly exceed the best fixed weight on Studies B, F, and L, the data its own thresholds were tuned against (fused selection correctness, gated minus best-fixed: gpt2, +0.0003; ngram5_kn, -0.0031), and did not exceed it on Study N (net correct gain, gated minus best-fixed: gpt2, -0.0128, 95% CI, -0.0271 to 0.0000; ngram5_kn, -0.0106, 95% CI, -0.0212 to -0.0021; eTable 8).

#### S1.7 Additional checks

Four additional checks supplement the principal comparison; none of them read Study N a second time. Three of them, the oracle tie structure, the context-grouped oracle-structure check, and the gate component-model characterization, are dev-only, computed on Studies B, F, and L alone. The fourth, the frozen single policy across outcomes, instead re-uses the original rescue analysis's own four-study population, as explained in that paragraph below.

**Oracle tie structure.** The grid oracle (S1.3) breaks ties among equally fused-correct beta values by whichever value gives the fused posterior the highest probability mass on the true target symbol, then by row order. Across the 2,903 dev-only selections, the number of equally-correct beta values per selection was, under gpt2: none for 544 selections, exactly one for 65, two for 90, three for 131, four for 235, and all five for 1,838; a unique oracle-optimal beta existed for 2.24% of selections, a tie (two or more equally correct betas) for 79.02%, and no correct beta at all for 18.74%. Under ngram5_kn: none for 409 selections, exactly one for 67, two for 60, three for 77, four for 114, and all five for 2,176; a unique oracle-optimal beta existed for 2.31%, a tie for 83.60%, and no correct beta at all for 14.09%. Ties, not unique optima, are therefore the typical case for this oracle under both priors (eTable9).

**Frozen single policy across outcomes.** The original fixed-versus-adaptive comparison (S1.3) selected a policy separately for each outcome under test. As an additional check, a single policy per prior was instead selected using only fused selection correctness, then applied unchanged, without refitting, to all four outcomes. This check runs on the rescue analysis's own population, all four studies (B, F, L, and N) under that analysis's own participant-grouped cross-validation, rather than on the dev-only subset the other three checks use. Re-running it is not a second read of the Study N holdout: the rescue analysis is part of the original Aims 1 to 4 comparison, it predates the joint arbitration gate, and it has no holdout concept of its own. The holdout belongs to the frozen-gate evaluation (S1.6), which this check neither invokes nor retunes. Under gpt2, the fixed and adaptive rates from this single frozen policy matched exactly for prior capture (0.0190 versus 0.0190), phantom agreement (0.0382 versus 0.0382), fused selection correctness (0.7373 versus 0.7373), and net correct gain (0.0193 versus 0.0193), each difference 0.0000 with a 95% confidence interval of approximately -0.0009 to 0.0009. Under ngram5_kn, the same exact match held for prior capture (0.0151 versus 0.0151), phantom agreement (0.1026 versus 0.1026), fused selection correctness (0.8055 versus 0.8055), and net correct gain (0.0875 versus 0.0875), each difference 0.0000 with a 95% confidence interval of 0.0000 to 0.0000. Freezing the policy on a single outcome did not manufacture an adaptive advantage that per-outcome selection alone had produced. The pooled rates coincided to full floating-point precision under both priors, but by two different routes. Under ngram5_kn the tuning step chose only fully degenerate anchor pairs (0.25 with 0.25, and 0.5 with 0.5), each of which holds the weight constant, so the tuned-adaptive arm reproduces the fixed-beta control selection by selection; that row-for-row identity is why its confidence interval has exactly zero width. Under gpt2 some folds instead chose a near-degenerate pair (0.9 with 1.1) alongside the degenerate 1.0 with 1.0 chosen in the others, so in those folds the adaptive arm applies a continuous per-selection weight and does not reproduce the control selection by selection. It changes which selections come out correct without changing how many, leaving the pooled rates identical but the confidence interval nonzero. Either way the result is the same degenerate or near-degenerate solution already reported for fused selection correctness and net correct gain in the fair-comparison search (S1.3, eTable6).

**Context-grouped oracle-structure check.** The oracle-structure check (S1.6) grouped cross-validation folds by participant; because the 2,903 dev-only selections span only 885 unique linguistic contexts, a repeated context could in principle fall on both sides of a participant-grouped fold. A second check grouped folds by linguistic context instead, so that no context crossed a fold, using the same 2,903 selections. The context-grouped macro one-vs-rest AUROC (area under the receiver operating characteristic curve) was 0.792 for gpt2, matching the participant-grouped 0.792 reported in S1.6 and eTable8 (rounded there to 0.79), and 0.812 for ngram5_kn against a participant-grouped 0.813 (rounded there to 0.81), a difference of 0.001 (eTable9). Two nuisance structures, participant identity and linguistic context, were therefore each controlled for in turn, and neither changed the result. Neither run controls both at once: the participant-grouped folds leave repeated contexts free to cross, and the context-grouped folds leave participants free to cross. This is evidence against either form of leakage on its own accounting for the signal, not proof that the signal is free of leakage of every kind.

**Gate component-model characterization.** The gate's two component classifiers, predicting phantom agreement and prior capture from the joint feature set (S1.6), were characterized directly on the same 2,903 dev-only selections (eTable9). Under gpt2, the phantom-agreement classifier reached an area under the receiver operating characteristic curve (AUROC) of 0.9410 and an average precision (AUPRC) of 0.3292 across 95 events, and the prior-capture classifier reached an AUROC of 0.8814 and an AUPRC of 0.0905 across 58 events. The net-benefit score V(x) (phantom-agreement probability minus prior-capture probability) ranged from -0.9994 to 0.7696, with quartiles -0.0017 (25th), -0.0001 (median), and 0.0024 (75th). The frozen gate's realized weight deviated from the best fixed weight on 363 of the 2,903 selections (12.50%); of those 363 deviations, 359 moved toward more language-model influence and 4 toward more neural influence. Both the gated and the best fixed weight lie on the fixed-weight grid, so each deviating selection's fused outcome under either one is already computed; comparing the two, 19 of the 363 deviations turned an incorrect selection correct, 18 turned a correct selection incorrect, and 326 left correctness unchanged, a net gain of one correct selection across all 2,903 (the +0.0003 dev-side figure in S1.6). Under ngram5_kn, the phantom-agreement classifier reached an AUROC of 0.9739 and an AUPRC of 0.7277 across 210 events, and the prior-capture classifier reached an AUROC of 0.8627 and an AUPRC of 0.0429 across 15 events; V(x) ranged from -0.0085 to 1.0, with quartiles 0.0012, 0.0045, and 0.0211. The gate deviated on 270 of the 2,903 selections (9.30%), all 270 toward more language-model influence and none toward more neural influence; of those 270, 5 turned an incorrect selection correct, 14 turned a correct selection incorrect, and 251 left correctness unchanged, a net loss of nine correct selections (the -0.0031 dev-side figure in S1.6). Both deviation rates are computed from the out-of-fold, cross-validated V(x), the same estimate the frozen thresholds were selected against, and not from the dev-wide refit V(x) that the Study N holdout evaluation itself used (S1.6).

#### S1.8 Actionability-aware counterfactual beta controller

Study N had already served as S1.6's one-shot holdout, so it was not reused as a second holdout for this materially different controller. Among the three source studies not yet used as any addendum's holdout, the smallest by selection count, Study B (18 participants, 846 selections), was designated as this controller's own holdout, fixed before this evaluation's own code touched real selection-level data and unrelated to any expected outcome; Study F, Study L, and Study N (2,527 selections, 29 participants) formed its development set, with Study N entering only as ordinary development data, weighted no differently from Study F or Study L. Every design choice below was made using only the development set; Study B was read exactly once, for the final evaluation, and never for label derivation, feature design, threshold selection, or model training.

**Labels.** For each selection and each fusion weight already in the existing grid {0.25, 0.5, 1, 2, 4}, the fused argmax symbol and fused correctness were already recorded (Fusion Arms; Methods), so no new fusion computation was needed. A selection was labelled actionable if its fused argmax symbol differed across at least two of the five weights, and non-actionable otherwise: 935 of 2,527 development-set selections (37.00%) were actionable under gpt2, and 507 of 2,527 (20.06%) under ngram5_kn.

**Stage 1 (actionability classifier).** An out-of-fold, participant-grouped (5-fold) logistic regression, with per-fold feature standardization fit on the training fold only, predicted the actionability label from the same fifteen-feature joint neural-and-language-model set used by the gate in S1.6. Out-of-fold performance on the development set: area under the receiver operating characteristic curve (AUROC), 0.9192, and average precision (AUPRC), 0.8344, for gpt2 (935 actionable of 2,527); AUROC, 0.9368, and AUPRC, 0.7504, for ngram5_kn (507 actionable of 2,527).

**Stage 2 (counterfactual correctness classifiers).** For each of the five fusion weights, a separate out-of-fold, participant-grouped logistic regression, with the same per-fold standardization, was fit on all 2,527 development-set selections (not only the actionable subset, since more training data only helps a model that is consulted only on actionable trials) to predict fused correctness at that weight from the same joint feature set. Out-of-fold AUROC (AUPRC) by weight, under gpt2: 0.25, 0.8411 (0.8800); 0.5, 0.8391 (0.9051); 1.0, 0.8377 (0.9112); 2.0, 0.8414 (0.9118); 4.0, 0.8481 (0.9128). Under ngram5_kn: 0.25, 0.8422 (0.9506); 0.5, 0.8503 (0.9541); 1.0, 0.8603 (0.9538); 2.0, 0.8674 (0.9524); 4.0, 0.8945 (0.9569).

**Combining rule and tau selection.** The controller defaulted every selection to its prior's own development-wide best fixed weight (chosen from the same five-value grid to maximize mean fused correctness on the full development set: 1.0 for gpt2, 0.5 for ngram5_kn) and consulted the Stage 2 models only when the Stage 1 probability exceeded a decision threshold tau, selecting the weight with the highest Stage 2 probability among the five. Tau was grid-searched out-of-fold on the development set over {0.3, 0.4, 0.5, 0.6, 0.7} to maximize development-side net correct gain, selecting tau = 0.7 for gpt2 and tau = 0.4 for ngram5_kn. On the development set, the resulting controller did not clearly exceed the best fixed weight for either prior (fused selection correctness, controller minus best-fixed: gpt2, -0.0028; ngram5_kn, +0.0051).

**Frozen evaluation.** The selected tau, best fixed weight, Stage 1 classifier, and all five Stage 2 classifiers were refit once on the entire development set (no further cross-validation) and applied, unchanged, to Study B. Neither prior beat best-fixed-weight fusion. Under gpt2, net correct gain was 0.0047 for the controller against 0.0083 for best-fixed weight, a difference of -0.0035 (95% CI, -0.0071 to 0.0000; the upper bound lands exactly at zero as a bootstrap percentile artifact of this 846-selection holdout, not a rounded value). Under ngram5_kn, net correct gain was 0.0201 for the controller against 0.0189 for best-fixed weight, a difference of +0.0012 (95% CI, -0.0033 to 0.0050). Both intervals include zero (eTable10).

#### S1.9 Decoded-history replay

Throughout the primary analysis and every supplementary check above, each selection's language-model prior was conditioned on the ground-truth copy-spelling prefix preceding it (Methods), which lets the language model see the correct linguistic history even on trials where a fusion arm would itself have decoded a different symbol. This check replaced that ground-truth prefix with a decoded-history replay: at each prior's own best-fixed weight, selected by the same rule S1.8 used but applied here across all four studies rather than S1.8's three-study development set (beta = 1.0 for gpt2, beta = 0.5 for ngram5_kn -- identical to S1.8's own values in this case), every selection's language-model prior was instead conditioned on the same fusion arm's own previously fused symbols, so an earlier decoding error propagates forward into the linguistic context exactly as it would for a real deployed speller. No new model fitting or threshold selection was performed; this check only rescored the existing best-fixed-weight arm under the alternative history, so no separate holdout was needed.

Decoded-history replay materially worsened both fused selection correctness and net correct gain relative to the ground-truth-prefix condition, for both priors, on all 3,373 selections. Under gpt2, fused selection correctness fell from 0.7373 (ground-truth prefix) to 0.7246 (decoded history) and net correct gain fell from 0.0193 to 0.0065, a difference of -0.0127 (95% CI, -0.0201 to -0.0068). Under ngram5_kn, fused selection correctness fell from 0.8076 to 0.7272 and net correct gain fell from 0.0895 to 0.0092, a difference of -0.0803 (95% CI, -0.1048 to -0.0566). Both intervals exclude zero (eTable11). The absolute degradation was about six times larger for ngram5_kn than for gpt2, and the relative loss of the prior's own net benefit was far larger still: net correct gain fell by 66% for gpt2 (retaining about a third of its ground-truth-prefix value) against 90% for ngram5_kn (retaining about a tenth), indicating the classical n-gram prior's net benefit depends much more heavily on the favorable, non-deployable linguistic context the ground-truth-prefix convention provides than the neural language model's does.

#### S1.10 Candidate-arbitration controller and selective error flagging

Study N had already served as S1.6's one-shot holdout and Study B had already served as S1.8's one-shot holdout, so neither, nor by extension any of the four source studies, could honestly be treated as a fresh, untouched holdout for this fourth redesign. Instead, all four studies were used in a nested leave-one-study-out design: for each study in turn, held out as the outer test fold, the fixed weight and the candidate-arbitration controller were both fit on the other three studies (with the controller's own inner cross-validation folds drawn only from those three), then frozen and applied once to the held-out study. Every selection received exactly one out-of-study prediction, from the single outer fold in which its own study was held out, and only the pooled four-fold predictions are compared against a nested-cross-validation best-fixed-weight comparator (the fixed weight chosen on each fold's own three held-in studies, applied uniformly to that fold's held-out study). This is disclosed throughout as a rigorous exploratory cross-study evaluation, not as independent or prospective validation, since this project's own earlier redesigns had already shaped every one of the four studies before this analysis began.

**Candidate collapse.** For each selection, the achievable candidate set was built as the union of the neural-only argmax and the fused argmax at every point on the fixed-weight grid {0.25, 0.5, 1, 2, 4}. Under gpt2, 66.85% of selections had exactly one achievable candidate, 27.63% had two, 5.16% had three, 0.33% had four, and 0.03% had five. Under ngram5_kn, 78.42% had one, 17.52% had two, 3.74% had three, and 0.33% had four. Most selections are therefore not actionable by any per-trial policy restricted to this grid, for either prior.

**Candidate ranker and combining rule.** For each achievable candidate, the exact beta at which it would tie the fixed-weight winner was computed from the linear fused log-score S(beta) = beta * log(P_neural) + log(P_lm), giving beta*_jk = -(log P_lm(j) - log P_lm(k)) / (log P_neural(j) - log P_neural(k)) for any two candidates j and k; this analytic formula was verified against a dense brute-force beta scan before use. A participant-grouped, 5-fold logistic regression, with per-fold feature standardization, was fit to predict whether each candidate is the true target from five features: the candidate's own neural and language-model posterior probabilities, its signed boundary distance to the fixed-weight winner, the width of the beta interval over which the fixed-weight winner remains the winner against every other symbol, and the log-odds between the candidate's neural and language-model log-probabilities. The combining rule defaulted every selection to the fixed-weight winner and overrode it only when the ranker's probability for the best alternative candidate exceeded the fixed winner's own probability by more than a margin delta (grid-searched out-of-fold over {0.0, 0.05, 0.1, 0.15, 0.2, 0.3} on each fold's own three held-in studies to maximize development-side correctness).

**Frozen nested-cross-validation evaluation.** Neither prior beat nested-cross-validation best-fixed-weight fusion. Under gpt2 (3,373 selections pooled across all four outer folds), overall correctness was 0.7350, a difference in net correct gain of -0.00237 (95% CI, -0.0085 to 0.0031) relative to the nested-cross-validation best-fixed comparator; of 166 total overrides, 42 helped, 50 hurt, and 74 had no effect on correctness, 4.74% of the fixed comparator's own errors were correctable, and the oracle-headroom-recovered fraction was -0.0457. Under ngram5_kn, overall correctness was 0.7904, a difference in net correct gain of -0.00949 (95% CI, -0.0148 to -0.0042); of 84 total overrides, 15 helped, 47 hurt, and 22 had no effect, 2.22% of the fixed comparator's own errors were correctable, and the oracle-headroom-recovered fraction was -0.2424 (eTable12). Phantom agreement (a correct fused output the neural evidence alone would not have produced) occurred in 2.34% of gpt2 selections and 9.10% of ngram5_kn selections; prior capture (a correct neural reading overturned into an error) occurred in 0.65% and 1.87%, respectively.

**Selective error flagging.** The candidate ranker's own estimated probability that the fixed winner was the correct symbol, p_hat(fixed winner), already computed for every selection as part of the combining rule above, was reused as a confidence score for flagging rather than for overriding. This probability is continuous and near-monotonically associated with the fixed comparator's own correctness (decile accuracy ranged from approximately 30% in the least-confident tenth of selections to 92% in the most-confident tenth, under gpt2; approximately 41% to 89% under ngram5_kn, with an intermediate decile reaching 98% before the top decile's own small-bin noise brought it back down to 89%). A risk-coverage curve, ranking confidence within each outer fold's own held-out study before pooling across the four studies (avoiding a cross-fold calibration-mismatch that a single global ranking of raw fold-specific probabilities would risk; four outer-fold logistic regressions are not comparable on a shared numeric scale without this), retained the highest-confidence fraction of selections at coverage levels from 100% down to 70% (5-point steps) and flagged the remainder. At 5% flagged, this flagged 13.2% of the fixed comparator's own errors under gpt2 (a 2.64-fold enrichment over the proportionate 5% baseline; 95% CI, 2.07 to 3.57) and 14.7% under ngram5_kn (a 2.93-fold enrichment; 95% CI, 2.24 to 4.09); at 10% flagged, it flagged 26.2% of errors under gpt2 (2.61-fold; 95% CI, 2.10 to 3.48) and 30.1% under ngram5_kn (3.00-fold; 95% CI, 2.42 to 3.94). This disproportionate error-flagging held in the same direction for both priors at both flagged fractions and was not driven by any single source study (per-study enrichment ranged 1.88- to 4.24-fold under gpt2 and 2.41- to 4.22-fold under ngram5_kn, eTable13).

Whether this signal requires the candidate-arbitration ranker at all, rather than a confidence score already available from fixed-weight fusion with no additional model fitting, is a separate question the ranker's own byproduct status does not answer. S1.11 benchmarks the ranker's confidence against seven confidence signals that require no fitted ranker; the ranker's confidence is not better than the simplest of them, and is significantly worse specifically on enrichment for both priors. The disproportionate-flagging capability itself is real, but it is carried by the fixed system's own posterior, not by the candidate-arbitration controller's added complexity (eTable13, eTable14).

#### S1.11 Benchmarking the error-flagging confidence signal against trivial baselines

The candidate ranker's confidence, p_hat(fixed winner), is one of several signals already available at inference time without fitting any additional model: the fixed-fused posterior's own top-1 probability, margin, and entropy (computed once the fixed weight is applied, S1.1); the neural posterior's own top-1 probability and margin (available before any language-model fusion); the trial-level reliability estimate, ĝ (Methods); and the Jensen-Shannon divergence between the neural and language-model posteriors. All eight signals were computed for every selection and compared at identifying the fixed comparator's own errors, using the same within-study-ranked, four-fold-pooled risk-coverage design as the ranker's own headline result above (S1.10), so no signal is advantaged by a different ranking convention.

Two comparisons were run. First, a participant-cluster bootstrap 95% confidence interval for each signal's own error-fraction-captured and enrichment at 5% and 10% flagged (eTable13); overlapping intervals do not establish equivalence when signals are computed on the same selections, since their per-selection outcomes are correlated, so a second, paired-difference participant-cluster bootstrap resampled the same participants for both signals within each bootstrap iteration and computed the ranker's own metric minus each baseline's, directly testing whether the ranker's confidence outperforms each alternative on the same resampled data (eTable14). A confidence interval excluding zero in this paired test is a materially stronger claim than a pair of overlapping unpaired intervals would be.

The ranker's confidence did not exceed any of the seven baseline signals on either metric at either coverage level for either prior (eTable14, all point estimates zero or negative). The paired difference against the fixed-fused posterior's own top-1 probability specifically, the single simplest available signal, excluded zero on enrichment for both priors (gpt2, difference, -0.428; 95% CI, -0.789 to -0.171; ngram5_kn, difference, -0.887; 95% CI, -1.470 to -0.489); the ranker's confidence was also significantly worse than the fixed-fused posterior's own entropy on enrichment for both priors. Two of seven comparisons ran in the opposite direction: the ranker's confidence was significantly better than the neural-language-model divergence signal (the weakest of the eight throughout) and, for ngram5_kn only, than the raw neural posterior's own margin at 10% flagged. No paired comparison found the ranker's confidence significantly better than any of the three fixed-fused-posterior signals on any metric, at any coverage, for either prior.

Error-fraction-captured and enrichment are not two independent lines of evidence: at a fixed coverage level every signal flags the identical number of selections by construction, so enrichment equals error-fraction-captured multiplied by a constant (selections divided by number flagged) that does not vary by signal, and the two columns of eTable14 differ only in how each metric's bootstrap distribution handles that shared underlying quantity. A calibration check simulating this study's exact design (3,373 selections, 47 participants, 4 studies, two equally informative correlated signals with a true difference of zero) found the enrichment interval's nominal 5% false-positive rate held closely (5.8% observed over 600 replicates) while the error-fraction-captured interval over-covered (2.3%), consistent with that count-based statistic's coarser lattice of achievable values. Enrichment is accordingly the metric this study treats as primary; the sparser significance calls on error-fraction-captured reflect that discreteness, not a weaker or contradictory effect.

None of the 56 signal-by-prior-by-coverage-by-metric comparisons in eTable14 was corrected for multiple comparisons, matching this study's existing disclosed convention for other uncorrected descriptive sweeps (S1.4). The two headline comparisons against the fixed-fused posterior's own top-1 probability sit far from the significance boundary for both priors; a handful of comparisons closer to the boundary (full values in eTable14) should be read as suggestive rather than confirmed. The bootstrap also resamples participants around each signal's already-fixed value rather than refitting the candidate ranker within each resample, so the ranker's own model-fitting uncertainty is not propagated into its interval, a limitation shared with this study's other bootstrap-based comparisons involving a fitted model (S1.6-S1.8) but not with the seven baseline signals, none of which require fitting.

Read together, the selective-error-flagging finding is genuine and safety-relevant: flagging the least-confident fraction of selections for review recovers a disproportionate share of the fixed comparator's own errors. The signal that does this work is the fixed-fused posterior already computed for fixed-weight fusion, available with no additional model fitting; the candidate-arbitration controller's own learned confidence, the byproduct this study originally reported the finding from, does not add value over that simpler signal and is measurably worse at identifying errors specifically (enrichment) for both language-model priors tested.

### S2 Supplementary Tables

Table eTable1. Per-arm outcome rates for the two primary-analysis priors. Mean rate of each outcome for every one of the four fusion arms (no-language-model floor, fixed-weight fusion, adaptive fusion, oracle), computed under a general-purpose large-language-model prior (gpt2) and a classical n-gram prior (ngram5_kn), each over all 3,373 selections.

| Prior | Arm | N | Fused correct | Neural correct | Prior capture | Phantom agreement | Net correct gain |
| --- | --- | --- | --- | --- | --- | --- | --- |
| gpt2 | adaptive | 3373 | 0.7305 | 0.7181 | 0.0246 | 0.0371 | 0.0125 |
| gpt2 | fixed | 3373 | 0.7373 | 0.7181 | 0.0190 | 0.0382 | 0.0193 |
| gpt2 | floor | 3373 | 0.7181 | 0.7181 | 0.0000 | 0.0000 | 0.0000 |
| gpt2 | oracle | 3373 | 0.7892 | 0.7181 | 0.0033 | 0.0744 | 0.0712 |
| ngram5_kn | adaptive | 3373 | 0.7931 | 0.7181 | 0.0059 | 0.0809 | 0.0750 |
| ngram5_kn | fixed | 3373 | 0.7969 | 0.7181 | 0.0053 | 0.0842 | 0.0789 |
| ngram5_kn | floor | 3373 | 0.7181 | 0.7181 | 0.0000 | 0.0000 | 0.0000 |
| ngram5_kn | oracle | 3373 | 0.8390 | 0.7181 | 0.0003 | 0.1213 | 0.1210 |

Table eTable2. Bootstrap contrasts on the principal and secondary outcomes. Participant-cluster bootstrap point estimate and 95% confidence interval for each arm-to-arm difference reported in the main text, plus the oracle-to-fixed gap-closure fraction on fused selection correctness.

| Prior | Outcome | Contrast | Difference | Lower 95% CI | Upper 95% CI |
| --- | --- | --- | --- | --- | --- |
| gpt2 | Net correct gain | Adaptive vs fixed | -0.0068 | -0.0128 | -0.0012 |
| gpt2 | Net correct gain | Adaptive vs no-language-model floor | 0.0125 | 0.0012 | 0.0238 |
| gpt2 | Net correct gain | Oracle vs fixed | 0.0519 | 0.0382 | 0.0655 |
| gpt2 | Prior capture | Adaptive vs fixed | 0.0056 | 0.0015 | 0.0104 |
| gpt2 | Phantom agreement | Adaptive vs fixed | -0.0012 | -0.0049 | 0.0024 |
| gpt2 | Fused selection correctness | Adaptive vs fixed | -0.0068 | -0.0128 | -0.0012 |
| gpt2 | Fused selection correctness | Oracle-to-fixed gap closed by adaptive | -0.1314 |  |  |
| ngram5_kn | Net correct gain | Adaptive vs fixed | -0.0039 | -0.0096 | 0.0020 |
| ngram5_kn | Net correct gain | Adaptive vs no-language-model floor | 0.0750 | 0.0552 | 0.0965 |
| ngram5_kn | Net correct gain | Oracle vs fixed | 0.0421 | 0.0286 | 0.0563 |
| ngram5_kn | Prior capture | Adaptive vs fixed | 0.0006 | -0.0015 | 0.0033 |
| ngram5_kn | Phantom agreement | Adaptive vs fixed | -0.0033 | -0.0092 | 0.0033 |
| ngram5_kn | Fused selection correctness | Adaptive vs fixed | -0.0039 | -0.0096 | 0.0020 |
| ngram5_kn | Fused selection correctness | Oracle-to-fixed gap closed by adaptive | -0.0915 |  |  |

Table eTable3. Generalized estimating equation odds ratios for the three binary secondary outcomes. Each row contrasts one arm against fixed-weight fusion (the reference level), from a binomial generalized estimating equation clustered on participant; the no-language-model floor arm is omitted from the prior-capture and phantom-agreement rows because it is 0% by construction for both.

| Prior | Outcome | Arm contrast (vs fixed) | Odds ratio | Lower 95% CI | Upper 95% CI | P value |
| --- | --- | --- | --- | --- | --- | --- |
| gpt2 | Prior capture | adaptive | 1.304 | 1.094 | 1.555 | 0.0031 |
| gpt2 | Prior capture | oracle | 0.169 | 0.116 | 0.248 | <0.001 |
| gpt2 | Phantom agreement | adaptive | 0.968 | 0.878 | 1.067 | 0.5099 |
| gpt2 | Phantom agreement | oracle | 2.022 | 1.750 | 2.336 | <0.001 |
| gpt2 | Fused selection correctness | floor | 0.907 | 0.866 | 0.951 | <0.001 |
| gpt2 | Fused selection correctness | adaptive | 0.966 | 0.939 | 0.994 | 0.0162 |
| gpt2 | Fused selection correctness | oracle | 1.334 | 1.273 | 1.398 | <0.001 |
| ngram5_kn | Prior capture | adaptive | 1.112 | 0.727 | 1.700 | 0.6249 |
| ngram5_kn | Prior capture | oracle | 0.055 | 0.008 | 0.383 | 0.0034 |
| ngram5_kn | Phantom agreement | adaptive | 0.958 | 0.879 | 1.043 | 0.3239 |
| ngram5_kn | Phantom agreement | oracle | 1.501 | 1.371 | 1.642 | <0.001 |
| ngram5_kn | Fused selection correctness | floor | 0.649 | 0.609 | 0.692 | <0.001 |
| ngram5_kn | Fused selection correctness | adaptive | 0.977 | 0.943 | 1.011 | 0.1807 |
| ngram5_kn | Fused selection correctness | oracle | 1.328 | 1.264 | 1.396 | <0.001 |

Table eTable4. Ladder-wide consistency sweep across 24 language-model priors. Adaptive-minus-fixed difference in prior-capture rate (positive favors fixed-weight fusion, since adaptive then causes more harm) for each of the 24 non-null priors in the companion study's ladder, spanning three classical n-grams and 21 causal language models across ten families.

| Prior model | Adaptive minus fixed (prior capture) | Lower 95% CI | Upper 95% CI |
| --- | --- | --- | --- |
| EleutherAI/pythia-12b | 0.0009 | -0.0023 | 0.0044 |
| Qwen/Qwen2.5-1.5B | 0.0036 | -0.0008 | 0.0083 |
| Qwen/Qwen2.5-14B | 0.0027 | -0.0016 | 0.0072 |
| Qwen/Qwen2.5-32B | 0.0021 | -0.0025 | 0.0067 |
| Qwen/Qwen2.5-3B | -0.0009 | -0.0069 | 0.0050 |
| Qwen/Qwen3.5-27B | 0.0047 | 0.0003 | 0.0095 |
| Qwen/Qwen3.6-35B-A3B | 0.0047 | 0.0000 | 0.0097 |
| allenai/OLMo-2-1124-7B-Instruct | 0.0000 | -0.0050 | 0.0056 |
| deepseek-ai/DeepSeek-V2-Lite | 0.0003 | -0.0031 | 0.0041 |
| google/gemma-2-27b | 0.0021 | -0.0021 | 0.0065 |
| google/gemma-2-2b-it | -0.0050 | -0.0101 | 0.0003 |
| google/gemma-2-9b | -0.0059 | -0.0117 | 0.0003 |
| google/gemma-4-12b-it | 0.0003 | -0.0064 | 0.0066 |
| google/recurrentgemma-2b | 0.0047 | 0.0009 | 0.0090 |
| gpt2 | 0.0056 | 0.0015 | 0.0104 |
| gpt2-large | 0.0056 | 0.0014 | 0.0103 |
| meta-llama/Llama-3.1-8B-Instruct | 0.0018 | -0.0035 | 0.0064 |
| mistralai/Mistral-7B-v0.3 | 0.0012 | -0.0032 | 0.0057 |
| mistralai/Mixtral-8x7B-v0.1 | 0.0018 | -0.0017 | 0.0055 |
| ngram5 | 0.0018 | -0.0009 | 0.0045 |
| ngram5_kn | 0.0006 | -0.0015 | 0.0033 |
| ngram5_wiki_kn | 0.0015 | -0.0006 | 0.0038 |
| openai/gpt-oss-20b | 0.0044 | 0.0009 | 0.0083 |
| state-spaces/mamba-2.8b-hf | 0.0024 | -0.0015 | 0.0064 |

Table eTable4b. Ladder-wide consistency sweep across 24 language-model priors, redesigned principal endpoint. Same design as eTable4, on fused selection correctness (the outcome fused_correct, whose difference is numerically identical to a difference in net correct gain within a fixed row set, since neural correctness does not vary across arms; see Methods). Fused selection correctness is a BENEFIT outcome (higher is better), the opposite direction-sense from prior_capture: here positive values favor adaptive fusion and negative values favor fixed-weight fusion.

| Prior model | Adaptive minus fixed (fused selection correctness) | Lower 95% CI | Upper 95% CI |
| --- | --- | --- | --- |
| EleutherAI/pythia-12b | -0.0062 | -0.0113 | -0.0012 |
| Qwen/Qwen2.5-1.5B | -0.0077 | -0.0130 | -0.0025 |
| Qwen/Qwen2.5-14B | -0.0086 | -0.0148 | -0.0029 |
| Qwen/Qwen2.5-32B | -0.0056 | -0.0114 | 0.0003 |
| Qwen/Qwen2.5-3B | -0.0050 | -0.0113 | 0.0019 |
| Qwen/Qwen3.5-27B | -0.0077 | -0.0127 | -0.0030 |
| Qwen/Qwen3.6-35B-A3B | -0.0113 | -0.0196 | -0.0038 |
| allenai/OLMo-2-1124-7B-Instruct | -0.0086 | -0.0166 | -0.0013 |
| deepseek-ai/DeepSeek-V2-Lite | -0.0080 | -0.0143 | -0.0016 |
| google/gemma-2-27b | -0.0021 | -0.0076 | 0.0036 |
| google/gemma-2-2b-it | -0.0015 | -0.0087 | 0.0047 |
| google/gemma-2-9b | -0.0042 | -0.0105 | 0.0020 |
| google/gemma-4-12b-it | -0.0042 | -0.0124 | 0.0043 |
| google/recurrentgemma-2b | -0.0098 | -0.0161 | -0.0039 |
| gpt2 | -0.0068 | -0.0128 | -0.0012 |
| gpt2-large | -0.0107 | -0.0166 | -0.0050 |
| meta-llama/Llama-3.1-8B-Instruct | -0.0065 | -0.0137 | 0.0006 |
| mistralai/Mistral-7B-v0.3 | -0.0044 | -0.0127 | 0.0024 |
| mistralai/Mixtral-8x7B-v0.1 | -0.0050 | -0.0110 | -0.0003 |
| ngram5 | -0.0083 | -0.0137 | -0.0020 |
| ngram5_kn | -0.0039 | -0.0096 | 0.0020 |
| ngram5_wiki_kn | -0.0077 | -0.0135 | -0.0019 |
| openai/gpt-oss-20b | -0.0056 | -0.0108 | -0.0006 |
| state-spaces/mamba-2.8b-hf | -0.0059 | -0.0121 | 0.0000 |

Table eTable5. Leave-one-study-out transportability. Adaptive-minus-fixed difference in prior-capture rate (positive favors fixed-weight fusion) when the reliability estimator is fit on three of the four source studies and evaluated on the held-out study, repeated for both primary-analysis priors and all four held-out choices.

| Prior | Held-out study | Adaptive minus fixed (prior capture) | Lower 95% CI | Upper 95% CI |
| --- | --- | --- | --- | --- |
| gpt2 | StudyB | 0.0047 | 0.0000 | 0.0089 |
| gpt2 | StudyF | 0.0009 | -0.0037 | 0.0057 |
| gpt2 | StudyL | 0.0141 | 0.0000 | 0.0293 |
| gpt2 | StudyN | 0.0064 | 0.0021 | 0.0126 |
| ngram5_kn | StudyB | 0.0000 | 0.0000 | 0.0000 |
| ngram5_kn | StudyF | -0.0019 | -0.0046 | 0.0000 |
| ngram5_kn | StudyL | 0.0051 | -0.0010 | 0.0131 |
| ngram5_kn | StudyN | -0.0021 | -0.0064 | 0.0000 |

Table eTable5b. Leave-one-study-out transportability, redesigned principal endpoint. Same design as eTable5, on fused selection correctness. As in eTable4b, positive values favor adaptive fusion and negative values favor fixed-weight fusion (the opposite direction-sense from eTable5's prior-capture convention).

| Prior | Held-out study | Adaptive minus fixed (fused selection correctness) | Lower 95% CI | Upper 95% CI |
| --- | --- | --- | --- | --- |
| gpt2 | StudyB | -0.0024 | -0.0072 | 0.0028 |
| gpt2 | StudyF | -0.0084 | -0.0192 | 0.0009 |
| gpt2 | StudyL | -0.0111 | -0.0233 | 0.0010 |
| gpt2 | StudyN | -0.0085 | -0.0216 | 0.0043 |
| ngram5_kn | StudyB | -0.0047 | -0.0165 | 0.0016 |
| ngram5_kn | StudyF | -0.0122 | -0.0237 | -0.0019 |
| ngram5_kn | StudyL | 0.0000 | -0.0121 | 0.0152 |
| ngram5_kn | StudyN | 0.0064 | -0.0043 | 0.0210 |

Table eTable6. Fair-comparison (rescue) results. Best-fixed-beta control versus tuned-adaptive arm, both chosen out-of-fold from a search space that fully contains the fixed control's own options (S1.3), for prior capture, fused selection correctness, and net correct gain, under both primary-analysis priors. The chosen fixed betas and chosen adaptive anchors are listed per prior and outcome.

| Prior | Outcome | Best-fixed rate | Tuned-adaptive rate | Difference | Lower 95% CI | Upper 95% CI | Chosen fixed betas | Chosen anchors |
| --- | --- | --- | --- | --- | --- | --- | --- | --- |
| gpt2 | Prior capture | 0.0033 | 0.0033 | 0.000000 | 0.000000 | 0.000000 | [4.0] | [[4.0, 4.0]] |
| gpt2 | Fused selection correctness | 0.7373 | 0.7373 | 0.000000 | -0.000830 | 0.000858 | [1.0] | [[0.9, 1.1], [1.0, 1.0]] |
| gpt2 | Net correct gain | 0.0193 | 0.0193 | 0.000000 | -0.000830 | 0.000858 |  |  |
| ngram5_kn | Prior capture | 0.0003 | 0.0003 | 0.000000 | 0.000000 | 0.000000 | [4.0] | [[4.0, 4.0]] |
| ngram5_kn | Fused selection correctness | 0.8055 | 0.8055 | 0.000000 | 0.000000 | 0.000000 | [0.25, 0.5] | [[0.25, 0.25], [0.5, 0.5]] |
| ngram5_kn | Net correct gain | 0.0875 | 0.0875 | 0.000000 | 0.000000 | 0.000000 |  |  |

Table eTable7. Benjamini-Hochberg-adjusted p values for the adaptive-versus-fixed contrast on the three binary secondary outcomes, within the family of six tests (three outcomes times two priors).

| Prior | Outcome | Contrast | Raw p | BH-adjusted p |
| --- | --- | --- | --- | --- |
| gpt2 | Prior capture | Adaptive vs fixed | 0.0031 | 0.0186 |
| gpt2 | Phantom agreement | Adaptive vs fixed | 0.5099 | 0.6118 |
| gpt2 | Fused selection correctness | Adaptive vs fixed | 0.0162 | 0.0487 |
| ngram5_kn | Prior capture | Adaptive vs fixed | 0.6249 | 0.6249 |
| ngram5_kn | Phantom agreement | Adaptive vs fixed | 0.3239 | 0.4858 |
| ngram5_kn | Fused selection correctness | Adaptive vs fixed | 0.1807 | 0.3614 |

Table eTable8. Joint neural-and-language-model gate (supplementary analysis, S1.6). Calibration check repeats the principal four-arm comparison with both posteriors temperature-calibrated, on Studies B, F, and L. Dev-side gate check compares the frozen gate against the best fixed weight on the same three studies its thresholds were tuned against. Holdout evaluation is the single evaluation on Study N, read once.

| Analysis | Prior | Comparison | Point estimate | Lower 95% CI | Upper 95% CI |
| --- | --- | --- | --- | --- | --- |
| Calibration check (net correct gain) | gpt2 | Adaptive-calibrated vs fixed | -0.0103 | -0.0219 | -0.0007 |
| Calibration check (net correct gain) | ngram5_kn | Adaptive-calibrated vs fixed | +0.0059 | -0.0040 | +0.0172 |
| Oracle-structure check (macro OvR AUROC) | gpt2 | Joint features vs grid-oracle beta | 0.79 |  |  |
| Oracle-structure check (macro OvR AUROC) | ngram5_kn | Joint features vs grid-oracle beta | 0.81 |  |  |
| Dev-side gate check (fused selection correctness) | gpt2 | Gated vs best-fixed | +0.0003 |  |  |
| Dev-side gate check (fused selection correctness) | ngram5_kn | Gated vs best-fixed | -0.0031 |  |  |
| Holdout evaluation, Study N (net correct gain) | gpt2 | Gated vs best-fixed | -0.0128 | -0.0271 | 0.0000 |
| Holdout evaluation, Study N (net correct gain) | ngram5_kn | Gated vs best-fixed | -0.0106 | -0.0212 | -0.0021 |

Blank CI cells are point comparisons on a fixed row set (no participant-cluster bootstrap computed) or a discriminative-performance metric rather than a rate difference.

Table eTable9. Additional checks (S1.7). Oracle tie structure, context-grouped oracle-structure check, and gate component-model characterization, for both primary-analysis priors. The frozen-single-policy-across-outcomes check (S1.7) is not repeated here because every one of its values is an exact fixed-versus-adaptive tie, already shown in full in the S1.7 text.

| Check | Prior | Metric | Value |
| --- | --- | --- | --- |
| Oracle tie structure | gpt2 | Fraction with a unique oracle-optimal beta | 0.0224 |
| Oracle tie structure | ngram5_kn | Fraction with a unique oracle-optimal beta | 0.0231 |
| Context-grouped oracle structure | gpt2 | Macro OvR AUROC | 0.792 |
| Context-grouped oracle structure | ngram5_kn | Macro OvR AUROC | 0.812 |
| Gate component (phantom agreement) | gpt2 | AUROC | 0.9410 |
| Gate component (phantom agreement) | ngram5_kn | AUROC | 0.9739 |
| Gate component (prior capture) | gpt2 | AUROC | 0.8814 |
| Gate component (prior capture) | ngram5_kn | AUROC | 0.8627 |
| Gate deviation rate (dev) | gpt2 | Fraction deviating from best fixed | 0.1250 |
| Gate deviation rate (dev) | ngram5_kn | Fraction deviating from best fixed | 0.0930 |
| Gate deviation outcome (dev) | gpt2 | Deviations helped / hurt / no effect | 19 / 18 / 326 |
| Gate deviation outcome (dev) | ngram5_kn | Deviations helped / hurt / no effect | 5 / 14 / 251 |

Table eTable10. Actionability-aware counterfactual beta controller (S1.8). Development-set figures are out-of-fold, computed on Study F, Study L, and Study N (2,527 selections). Holdout figures are the single evaluation on Study B (18 participants, 846 selections), read once.

| Analysis | Prior | Comparison | Point estimate | Lower 95% CI | Upper 95% CI |
| --- | --- | --- | --- | --- | --- |
| Actionable rate (dev) | gpt2 | Fraction of selections flagged actionable | 0.3700 (935/2527) |  |  |
| Actionable rate (dev) | ngram5_kn | Fraction of selections flagged actionable | 0.2006 (507/2527) |  |  |
| Stage 1 discrimination (dev) | gpt2 | AUROC | 0.9192 |  |  |
| Stage 1 discrimination (dev) | gpt2 | AUPRC | 0.8344 |  |  |
| Stage 1 discrimination (dev) | ngram5_kn | AUROC | 0.9368 |  |  |
| Stage 1 discrimination (dev) | ngram5_kn | AUPRC | 0.7504 |  |  |
| Stage 2 discrimination at beta = 0.25 (dev) | gpt2 | AUROC (AUPRC) | 0.8411 (0.8800) |  |  |
| Stage 2 discrimination at beta = 0.5 (dev) | gpt2 | AUROC (AUPRC) | 0.8391 (0.9051) |  |  |
| Stage 2 discrimination at beta = 1.0 (dev) | gpt2 | AUROC (AUPRC) | 0.8377 (0.9112) |  |  |
| Stage 2 discrimination at beta = 2.0 (dev) | gpt2 | AUROC (AUPRC) | 0.8414 (0.9118) |  |  |
| Stage 2 discrimination at beta = 4.0 (dev) | gpt2 | AUROC (AUPRC) | 0.8481 (0.9128) |  |  |
| Stage 2 discrimination at beta = 0.25 (dev) | ngram5_kn | AUROC (AUPRC) | 0.8422 (0.9506) |  |  |
| Stage 2 discrimination at beta = 0.5 (dev) | ngram5_kn | AUROC (AUPRC) | 0.8503 (0.9541) |  |  |
| Stage 2 discrimination at beta = 1.0 (dev) | ngram5_kn | AUROC (AUPRC) | 0.8603 (0.9538) |  |  |
| Stage 2 discrimination at beta = 2.0 (dev) | ngram5_kn | AUROC (AUPRC) | 0.8674 (0.9524) |  |  |
| Stage 2 discrimination at beta = 4.0 (dev) | ngram5_kn | AUROC (AUPRC) | 0.8945 (0.9569) |  |  |
| Dev-side controller check (fused selection correctness) | gpt2 | Controller vs best-fixed | -0.0028 |  |  |
| Dev-side controller check (fused selection correctness) | ngram5_kn | Controller vs best-fixed | +0.0051 |  |  |
| Holdout evaluation, Study B (net correct gain) | gpt2 | Controller vs best-fixed | -0.0035 | -0.0071 | 0.0000 |
| Holdout evaluation, Study B (net correct gain) | ngram5_kn | Controller vs best-fixed | +0.0012 | -0.0033 | 0.0050 |

Blank CI cells are point comparisons on a fixed row set (no participant-cluster bootstrap computed) or a discriminative-performance metric rather than a rate difference. The holdout row's upper 95% CI bound for gpt2 lands exactly at 0.0000 as a bootstrap percentile artifact of the 846-selection holdout, not a rounded value.

Table eTable11. Decoded-history replay (S1.9). Fused selection correctness and net correct gain under the ground-truth copy-spelling prefix used throughout this study and under a decoded-history replay conditioning each prior on the same fusion arm's own previously decoded symbols, at each prior's own best-fixed weight (S1.8), across all 3,373 selections. For ngram5_kn this best-fixed weight is beta = 0.5, not the field-default beta = 1 "fixed" arm reported in eTable1, so the ground-truth-prefix row here is not the same number as eTable1's ngram5_kn/fixed row.

| Analysis | Prior | Comparison | Point estimate | Lower 95% CI | Upper 95% CI |
| --- | --- | --- | --- | --- | --- |
| Fused selection correctness | gpt2 | Ground-truth prefix | 0.7373 |  |  |
| Fused selection correctness | gpt2 | Decoded-history replay | 0.7246 |  |  |
| Fused selection correctness | ngram5_kn | Ground-truth prefix | 0.8076 |  |  |
| Fused selection correctness | ngram5_kn | Decoded-history replay | 0.7272 |  |  |
| Net correct gain | gpt2 | Ground-truth prefix | 0.0193 |  |  |
| Net correct gain | gpt2 | Decoded-history replay | 0.0065 |  |  |
| Net correct gain | gpt2 | Decoded-history minus ground-truth prefix | -0.0127 | -0.0201 | -0.0068 |
| Net correct gain | ngram5_kn | Ground-truth prefix | 0.0895 |  |  |
| Net correct gain | ngram5_kn | Decoded-history replay | 0.0092 |  |  |
| Net correct gain | ngram5_kn | Decoded-history minus ground-truth prefix | -0.0803 | -0.1048 | -0.0566 |

Blank CI cells are point rates on a fixed row set (no participant-cluster bootstrap computed per condition); only the decoded-minus-ground-truth-prefix contrast has a bootstrap 95% CI.

Table eTable12. Candidate-arbitration controller and selective error flagging (S1.10). Nested-cross-validation figures pool all four outer folds (3,373 selections total per prior). The net-correct-gain, override-outcome, and correctable-fraction rows were also computed for three further priors spanning this study's parameter range (Qwen2.5-1.5B, gemma-4-12b, Mixtral-8x7B), using the identical nested-leave-one-study-out procedure; the remaining rows (candidate collapse, overall correctness, oracle-headroom-recovered fraction, selective error flagging) were not re-run for these three and are reported only for gpt2/ngram5_kn. Selective-error-flagging rows report the fraction of the fixed comparator's own errors flagged at each flagged fraction of selections and the resulting disproportionality ratio (fraction of errors flagged divided by fraction flagged; 1.0 is proportionate, above 1.0 is disproportionate), using the within-study-ranked, four-fold-pooled design described in S1.10 and benchmarked against seven baseline confidence signals in S1.11 (eTable13, eTable14).

| Analysis | Prior | Comparison | Point estimate | Lower 95% CI | Upper 95% CI |
| --- | --- | --- | --- | --- | --- |
| Candidate collapse | gpt2 | 1 / 2 / 3 / 4 / 5 achievable candidates | 66.85% / 27.63% / 5.16% / 0.33% / 0.03% |  |  |
| Candidate collapse | ngram5_kn | 1 / 2 / 3 / 4 achievable candidates | 78.42% / 17.52% / 3.74% / 0.33% |  |  |
| Nested-CV overall correctness | gpt2 | Controller | 0.7350 |  |  |
| Nested-CV overall correctness | ngram5_kn | Controller | 0.7904 |  |  |
| Nested-CV net correct gain (controller vs nested-CV best-fixed) | gpt2 | Difference | -0.00237 | -0.0085 | 0.0031 |
| Nested-CV net correct gain (controller vs nested-CV best-fixed) | ngram5_kn | Difference | -0.00949 | -0.0148 | -0.0042 |
| Nested-CV net correct gain (controller vs nested-CV best-fixed) | Qwen2.5-1.5B (1.5B params) | Difference | -0.0036 | -0.0093 | 0.0020 |
| Nested-CV net correct gain (controller vs nested-CV best-fixed) | gemma-4-12b (12B params) | Difference | -0.0080 | -0.0148 | -0.0017 |
| Nested-CV net correct gain (controller vs nested-CV best-fixed) | Mixtral-8x7B (46.7B params) | Difference | -0.0033 | -0.0082 | 0.0011 |
| Override outcome (helped / hurt / no effect) | gpt2 | of 166 total overrides | 42 / 50 / 74 |  |  |
| Override outcome (helped / hurt / no effect) | ngram5_kn | of 84 total overrides | 15 / 47 / 22 |  |  |
| Override outcome (helped / hurt / no effect) | Qwen2.5-1.5B (1.5B params) | of 171 total overrides | 46 / 58 / 67 |  |  |
| Override outcome (helped / hurt / no effect) | gemma-4-12b (12B params) | of 173 total overrides | 36 / 63 / 74 |  |  |
| Override outcome (helped / hurt / no effect) | Mixtral-8x7B (46.7B params) | of 175 total overrides | 45 / 56 / 74 |  |  |
| Fraction of fixed comparator's own errors correctable by the controller | gpt2 | Controller | 0.0474 |  |  |
| Fraction of fixed comparator's own errors correctable by the controller | ngram5_kn | Controller | 0.0222 |  |  |
| Fraction of fixed comparator's own errors correctable by the controller | Qwen2.5-1.5B (1.5B params) | Controller | 0.0616 |  |  |
| Fraction of fixed comparator's own errors correctable by the controller | gemma-4-12b (12B params) | Controller | 0.0441 |  |  |
| Fraction of fixed comparator's own errors correctable by the controller | Mixtral-8x7B (46.7B params) | Controller | 0.0595 |  |  |
| Oracle-headroom-recovered fraction | gpt2 | Controller | -0.0457 |  |  |
| Oracle-headroom-recovered fraction | ngram5_kn | Controller | -0.2424 |  |  |
| Prior capture rate | gpt2 | Controller | 0.0065 |  |  |
| Prior capture rate | ngram5_kn | Controller | 0.0187 |  |  |
| Phantom agreement rate | gpt2 | Controller | 0.0234 |  |  |
| Phantom agreement rate | ngram5_kn | Controller | 0.0910 |  |  |
| Selective error flagging (candidate-arbitration ranker), 5% flagged | gpt2 | Fraction of errors flagged (disproportionality ratio) | 13.2% (2.64x) |  |  |
| Selective error flagging (candidate-arbitration ranker), 5% flagged | ngram5_kn | Fraction of errors flagged (disproportionality ratio) | 14.7% (2.93x) |  |  |
| Selective error flagging (candidate-arbitration ranker), 10% flagged | gpt2 | Fraction of errors flagged (disproportionality ratio) | 26.2% (2.61x) |  |  |
| Selective error flagging (candidate-arbitration ranker), 10% flagged | ngram5_kn | Fraction of errors flagged (disproportionality ratio) | 30.1% (3.00x) |  |  |

The candidate-collapse, nested-cross-validation overall-correctness, override-outcome, correctability, oracle-headroom, prior-capture, and phantom-agreement rows above have no participant-cluster bootstrap computed; they are nested-cross-validation pooled rates or ratios on a fixed row set, matching eTable9's and eTable10's convention for non-bootstrapped diagnostic figures. The net-correct-gain rows and the selective-error-flagging rows do have participant-cluster bootstrap 95% confidence intervals; the selective-error-flagging intervals are reported alongside a seven-signal baseline benchmark in eTable13 and eTable14.

Table eTable13. Selective-error-flagging confidence-signal benchmark (S1.11). Fraction of the fixed comparator's own errors flagged and the resulting enrichment (disproportionality ratio) for eight confidence signals, each ranked within its own held-out study before the four studies are pooled, at two flagged fractions of selections. Participant-cluster bootstrap 95% confidence intervals in brackets. The candidate-arbitration ranker (first row of each prior) is the signal this study originally reported the finding from; the remaining seven rows require no fitted ranker.

| Prior | Confidence signal | Flagged | Fraction of errors flagged [95% CI] | Enrichment [95% CI] |
| --- | --- | --- | --- | --- |
| gpt2 | Candidate-arbitration ranker (p_hat, fixed winner correct) | 5% | 13.2% [10.5, 16.5] | 2.64 [2.07, 3.57] |
| gpt2 | Candidate-arbitration ranker (p_hat, fixed winner correct) | 10% | 26.2% [21.7, 31.7] | 2.61 [2.10, 3.48] |
| gpt2 | Fixed-fused posterior, top-1 probability | 5% | 15.3% [11.6, 19.5] | 3.06 [2.41, 4.15] |
| gpt2 | Fixed-fused posterior, top-1 probability | 10% | 27.9% [23.3, 33.2] | 2.78 [2.24, 3.66] |
| gpt2 | Fixed-fused posterior, top-1-to-top-2 margin | 5% | 14.7% [11.7, 17.9] | 2.93 [2.31, 3.96] |
| gpt2 | Fixed-fused posterior, top-1-to-top-2 margin | 10% | 27.9% [23.7, 32.9] | 2.78 [2.25, 3.65] |
| gpt2 | Fixed-fused posterior, entropy (bits) | 5% | 15.0% [10.6, 20.1] | 3.00 [2.36, 4.08] |
| gpt2 | Fixed-fused posterior, entropy (bits) | 10% | 28.8% [23.1, 35.8] | 2.87 [2.31, 3.81] |
| gpt2 | Neural posterior, top-1 probability | 5% | 12.5% [8.7, 17.1] | 2.50 [1.97, 3.38] |
| gpt2 | Neural posterior, top-1 probability | 10% | 24.9% [19.9, 31.3] | 2.49 [2.02, 3.26] |
| gpt2 | Neural posterior, top-1-to-top-2 margin | 5% | 12.6% [9.5, 16.5] | 2.52 [2.03, 3.34] |
| gpt2 | Neural posterior, top-1-to-top-2 margin | 10% | 23.9% [18.8, 30.2] | 2.39 [1.94, 3.10] |
| gpt2 | Trial-level reliability estimate (ĝ) | 5% | 13.7% [8.7, 19.9] | 2.73 [2.16, 3.65] |
| gpt2 | Trial-level reliability estimate (ĝ) | 10% | 26.2% [19.5, 34.8] | 2.61 [2.10, 3.45] |
| gpt2 | Neural-language-model Jensen-Shannon divergence | 5% | 1.9% [1.1, 2.8] | 0.38 [0.21, 0.56] |
| gpt2 | Neural-language-model Jensen-Shannon divergence | 10% | 6.9% [5.1, 8.9] | 0.69 [0.52, 0.86] |
| ngram5_kn | Candidate-arbitration ranker (p_hat, fixed winner correct) | 5% | 14.7% [11.4, 18.6] | 2.93 [2.24, 4.09] |
| ngram5_kn | Candidate-arbitration ranker (p_hat, fixed winner correct) | 10% | 30.1% [24.4, 36.2] | 3.00 [2.42, 3.94] |
| ngram5_kn | Fixed-fused posterior, top-1 probability | 5% | 19.1% [14.5, 24.7] | 3.81 [3.01, 5.21] |
| ngram5_kn | Fixed-fused posterior, top-1 probability | 10% | 33.9% [27.8, 41.0] | 3.39 [2.72, 4.56] |
| ngram5_kn | Fixed-fused posterior, top-1-to-top-2 margin | 5% | 18.1% [13.2, 24.1] | 3.61 [2.86, 4.85] |
| ngram5_kn | Fixed-fused posterior, top-1-to-top-2 margin | 10% | 33.5% [27.9, 39.9] | 3.34 [2.66, 4.49] |
| ngram5_kn | Fixed-fused posterior, entropy (bits) | 5% | 17.5% [12.8, 22.8] | 3.49 [2.75, 4.76] |
| ngram5_kn | Fixed-fused posterior, entropy (bits) | 10% | 33.6% [27.4, 40.5] | 3.36 [2.67, 4.56] |
| ngram5_kn | Neural posterior, top-1 probability | 5% | 14.5% [9.9, 20.0] | 2.90 [2.30, 3.93] |
| ngram5_kn | Neural posterior, top-1 probability | 10% | 26.4% [20.0, 34.0] | 2.63 [2.15, 3.41] |
| ngram5_kn | Neural posterior, top-1-to-top-2 margin | 5% | 14.4% [10.4, 19.1] | 2.87 [2.32, 3.78] |
| ngram5_kn | Neural posterior, top-1-to-top-2 margin | 10% | 25.8% [19.9, 32.6] | 2.57 [2.11, 3.31] |
| ngram5_kn | Trial-level reliability estimate (ĝ) | 5% | 14.8% [9.7, 21.1] | 2.96 [2.33, 4.12] |
| ngram5_kn | Trial-level reliability estimate (ĝ) | 10% | 27.6% [20.2, 36.7] | 2.75 [2.24, 3.59] |
| ngram5_kn | Neural-language-model Jensen-Shannon divergence | 5% | 10.8% [8.1, 14.1] | 2.16 [1.54, 3.06] |
| ngram5_kn | Neural-language-model Jensen-Shannon divergence | 10% | 21.5% [16.7, 26.8] | 2.14 [1.67, 2.81] |

Table eTable14. Paired-difference participant-cluster bootstrap: candidate-arbitration ranker minus each baseline signal (S1.11). Same resampled participants applied to both signals within each bootstrap iteration; a negative difference favors the baseline. Bracketed intervals excluding zero are marked significant.

| Prior | Ranker minus | Flagged | Δ Fraction of errors flagged [95% CI] | Δ Enrichment [95% CI] |
| --- | --- | --- | --- | --- |
| gpt2 | Fixed-fused posterior, top-1 probability | 5% | -0.021 [-0.043, 0.000] | -0.428 [-0.789, -0.171], significant |
| gpt2 | Fixed-fused posterior, top-1 probability | 10% | -0.017 [-0.039, 0.008] | -0.169 [-0.357, -0.006], significant |
| gpt2 | Fixed-fused posterior, top-1-to-top-2 margin | 5% | -0.015 [-0.038, 0.008] | -0.293 [-0.666, 0.016] |
| gpt2 | Fixed-fused posterior, top-1-to-top-2 margin | 10% | -0.017 [-0.044, 0.013] | -0.169 [-0.361, 0.010] |
| gpt2 | Fixed-fused posterior, entropy (bits) | 5% | -0.018 [-0.044, 0.009] | -0.360 [-0.657, -0.157], significant |
| gpt2 | Fixed-fused posterior, entropy (bits) | 10% | -0.026 [-0.054, 0.005] | -0.259 [-0.440, -0.107], significant |
| gpt2 | Neural posterior, top-1 probability | 5% | 0.007 [-0.015, 0.027] | 0.135 [-0.080, 0.404] |
| gpt2 | Neural posterior, top-1 probability | 10% | 0.012 [-0.018, 0.044] | 0.124 [-0.011, 0.328] |
| gpt2 | Neural posterior, top-1-to-top-2 margin | 5% | 0.006 [-0.020, 0.027] | 0.113 [-0.138, 0.443] |
| gpt2 | Neural posterior, top-1-to-top-2 margin | 10% | 0.023 [-0.003, 0.051] | 0.225 [0.083, 0.489], significant (favors ranker) |
| gpt2 | Trial-level reliability estimate (ĝ) | 5% | -0.005 [-0.044, 0.030] | -0.090 [-0.420, 0.280] |
| gpt2 | Trial-level reliability estimate (ĝ) | 10% | 0.000 [-0.051, 0.048] | 0.000 [-0.128, 0.164] |
| gpt2 | Neural-language-model Jensen-Shannon divergence | 5% | 0.113 [0.081, 0.151], significant (favors ranker) | 2.253 [1.592, 3.290], significant (favors ranker) |
| gpt2 | Neural-language-model Jensen-Shannon divergence | 10% | 0.193 [0.140, 0.256], significant (favors ranker) | 1.926 [1.324, 2.881], significant (favors ranker) |
| ngram5_kn | Fixed-fused posterior, top-1 probability | 5% | -0.044 [-0.081, -0.008], significant | -0.887 [-1.470, -0.489], significant |
| ngram5_kn | Fixed-fused posterior, top-1 probability | 10% | -0.039 [-0.086, 0.004] | -0.384 [-0.785, -0.121], significant |
| ngram5_kn | Fixed-fused posterior, top-1-to-top-2 margin | 5% | -0.034 [-0.076, 0.007] | -0.680 [-1.190, -0.243], significant |
| ngram5_kn | Fixed-fused posterior, top-1-to-top-2 margin | 10% | -0.034 [-0.085, 0.011] | -0.340 [-0.734, -0.076], significant |
| ngram5_kn | Fixed-fused posterior, entropy (bits) | 5% | -0.028 [-0.067, 0.009] | -0.562 [-1.178, -0.015], significant |
| ngram5_kn | Fixed-fused posterior, entropy (bits) | 10% | -0.036 [-0.084, 0.010] | -0.355 [-0.749, -0.102], significant |
| ngram5_kn | Neural posterior, top-1 probability | 5% | 0.002 [-0.036, 0.036] | 0.030 [-0.468, 0.532] |
| ngram5_kn | Neural posterior, top-1 probability | 10% | 0.037 [-0.004, 0.079] | 0.370 [0.134, 0.738], significant (favors ranker) |
| ngram5_kn | Neural posterior, top-1-to-top-2 margin | 5% | 0.003 [-0.035, 0.040] | 0.059 [-0.487, 0.688] |
| ngram5_kn | Neural posterior, top-1-to-top-2 margin | 10% | 0.043 [0.005, 0.078], significant (favors ranker) | 0.429 [0.186, 0.815], significant (favors ranker) |
| ngram5_kn | Trial-level reliability estimate (ĝ) | 5% | -0.002 [-0.046, 0.040] | -0.030 [-0.678, 0.525] |
| ngram5_kn | Trial-level reliability estimate (ĝ) | 10% | 0.025 [-0.028, 0.073] | 0.251 [-0.010, 0.540] |
| ngram5_kn | Neural-language-model Jensen-Shannon divergence | 5% | 0.039 [-0.010, 0.088] | 0.769 [0.001, 1.635], significant (favors ranker) |
| ngram5_kn | Neural-language-model Jensen-Shannon divergence | 10% | 0.086 [0.000, 0.175] | 0.858 [0.319, 1.561], significant (favors ranker) |

Per-study enrichment for the candidate-arbitration ranker alone, at 5% flagged: gpt2 ranged 1.88-fold (Study N) to 4.24-fold (Study B); ngram5_kn ranged 2.41-fold (Study F) to 4.22-fold (Study B). At 10% flagged: gpt2 ranged 1.87-fold (Study N) to 4.28-fold (Study B); ngram5_kn ranged 2.39-fold (Study F) to 4.64-fold (Study B). No study drove the pooled result or reversed its direction for either prior.

### S3 Supplementary Figures

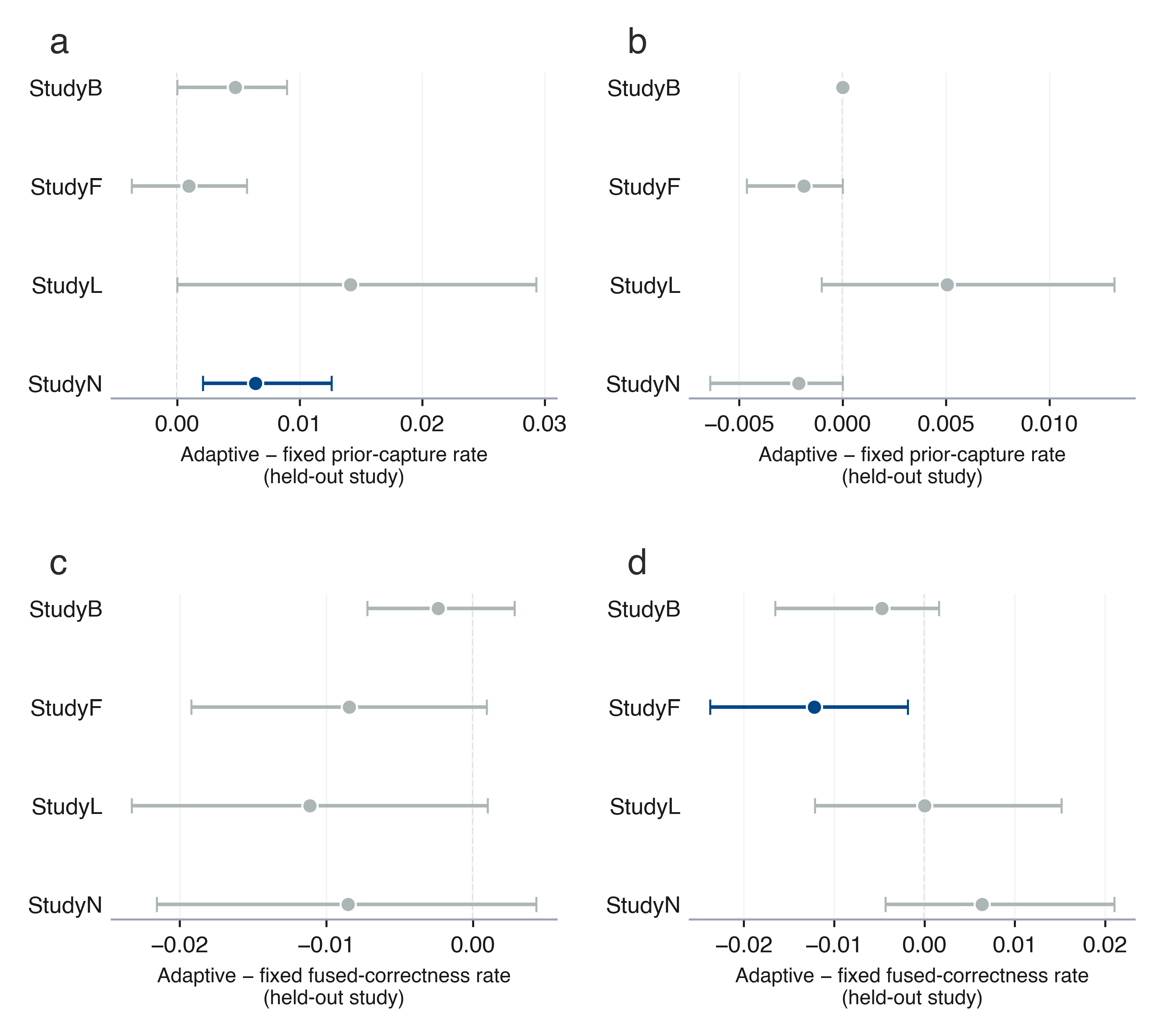

eFigure 1. Leave-one-study-out transportability of the main fixed-versus-adaptive contrast, for a general-purpose language-model prior (left column, gpt2) and a classical n-gram prior (right column, ngram5_kn), on prior capture (top row, panels a-b) and fused selection correctness (bottom row, panels c-d; eTable 5b). Points are the adaptive-minus-fixed difference for each held-out study; error bars are 95% confidence intervals; color marks whether the interval excludes zero.

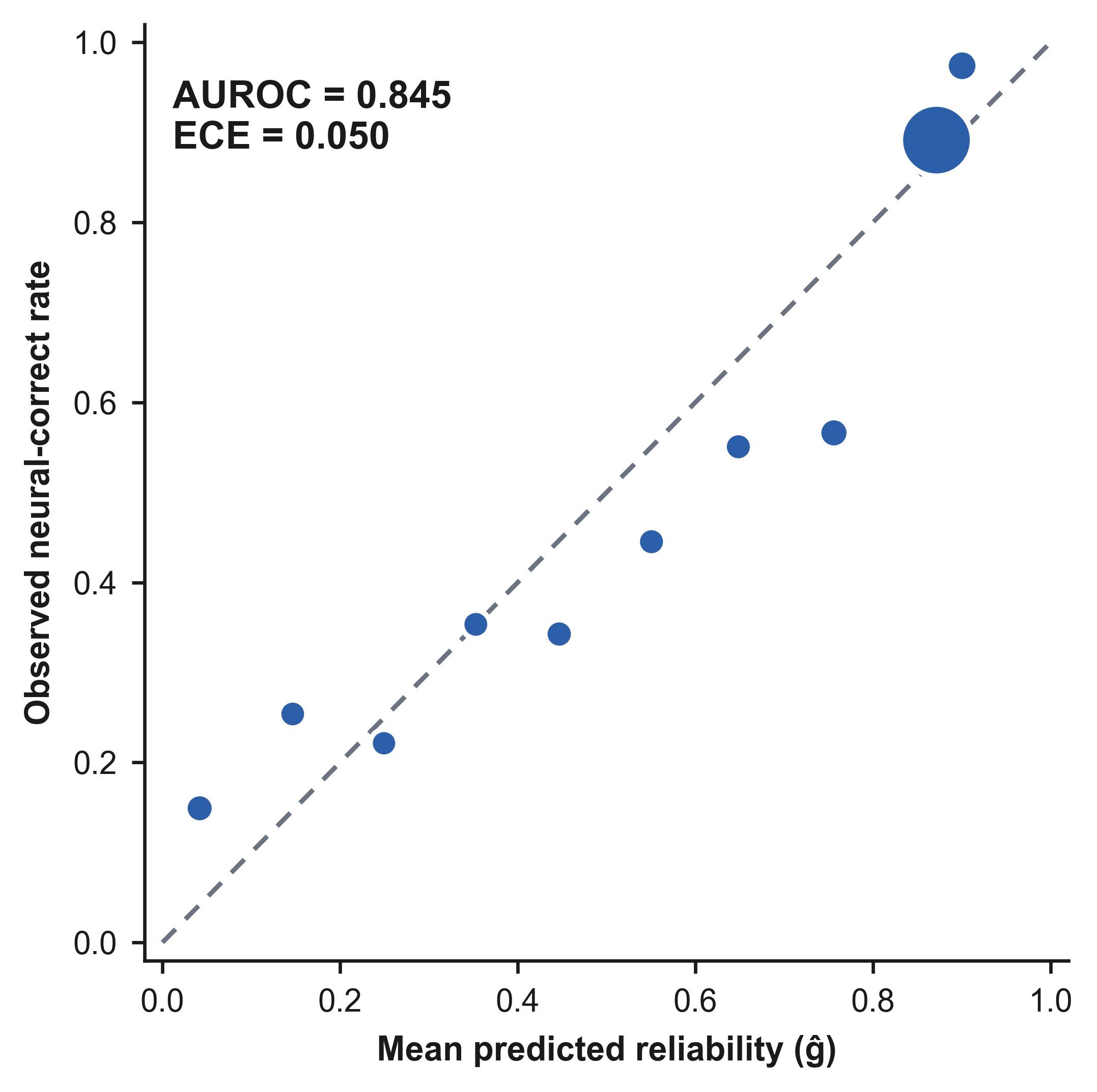

eFigure 2. The reliability estimate discriminates correct from incorrect neural selections well, but that signal does not by itself imply an actionable fusion decision (Results: A predictive reliability estimate with little to act on). Reliability diagram (observed frequency of a correct neural selection against the estimator's predicted probability, in ten equal-width bins); point size is proportional to the number of selections in that bin. Out-of-fold AUROC and expected calibration error are annotated; the diagonal marks perfect calibration.

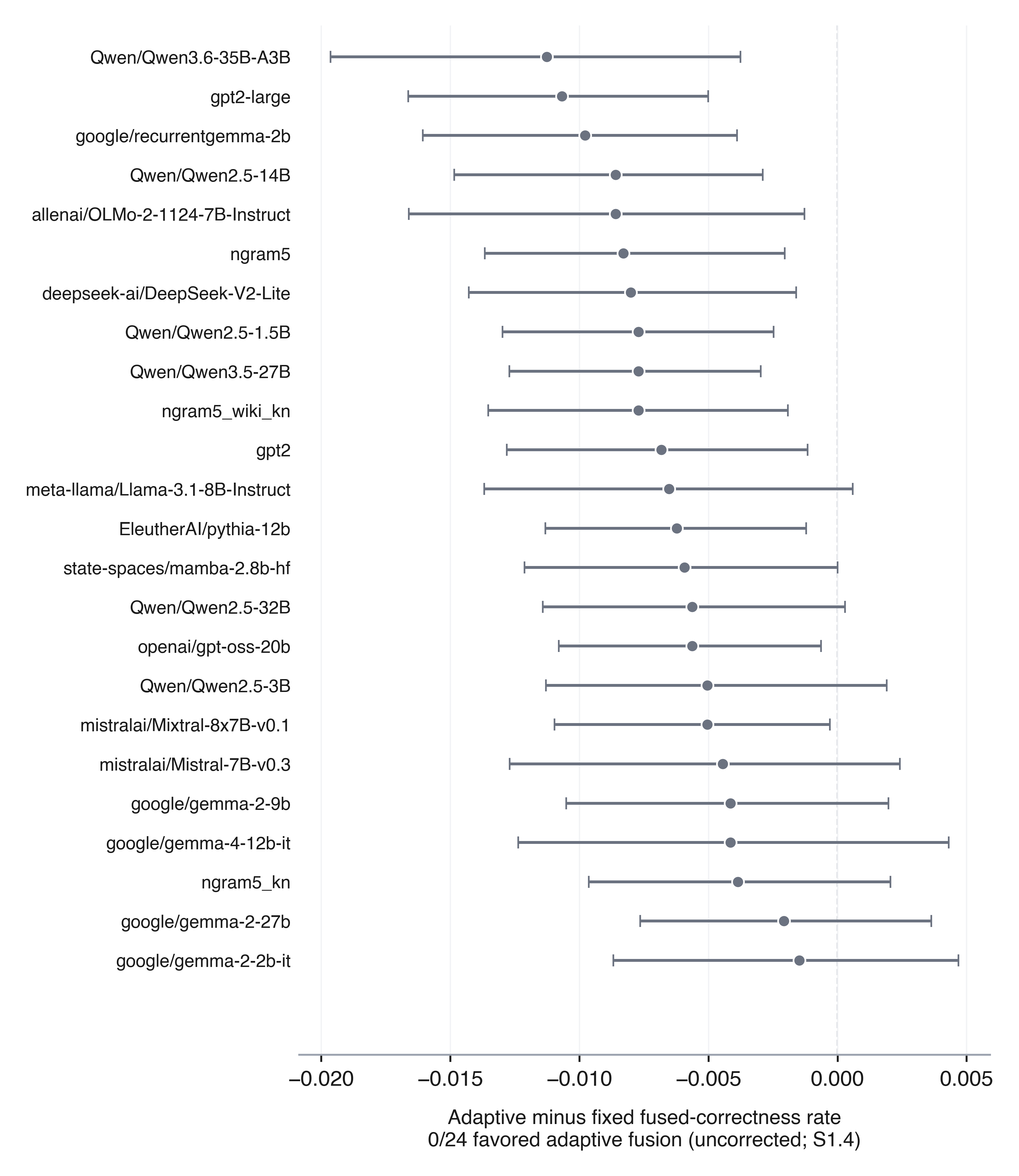

eFigure 3. Ladder-wide consistency sweep on fused selection correctness, 24 language-model priors (Results: ladder-wide consistency; eTable 4b). Adaptive-minus-fixed difference in fused-selection- correctness rate for each prior, sorted by effect size; error bars are 95% participant-cluster bootstrap confidence intervals, uncorrected for multiplicity across these 24 comparisons (S1.4). All points are drawn in one neutral color and the panel reports the point-estimate count directly, since color-coding by whether an individual CI excludes zero would invite reading this sweep as a per-model significance count rather than the descriptive consistency check it is.

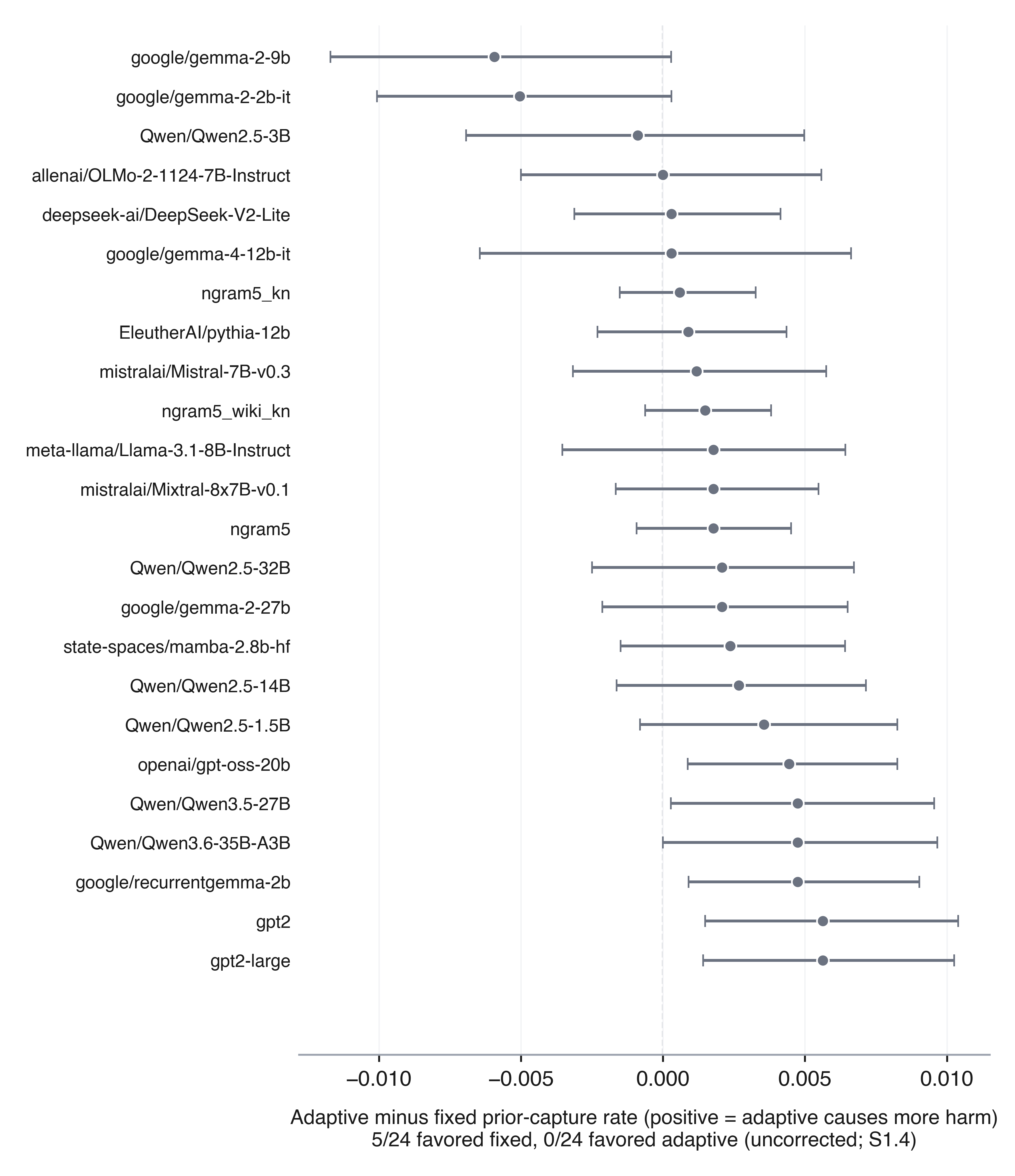

eFigure 4. Ladder-wide consistency sweep on prior capture, the same 24 language-model priors (Results: ladder-wide consistency; eTable 4). Adaptive-minus-fixed difference in prior-capture rate for each prior, sorted by effect size; positive values indicate adaptive fusion caused more harm. Points are the participant-cluster bootstrap point estimate; error bars are 95% confidence intervals. As in eFigure 3, all points are drawn in one neutral color and the panel reports the point-estimate counts directly, since these 24 comparisons are uncorrected for multiplicity (S1.4) and color-coding by whether an individual CI excludes zero would invite reading this sweep as a per-model significance count rather than the descriptive consistency check it is. The fair, matched-search-space comparison across all 22 evaluable priors is Figure 1 in the main text, not a supplementary figure.

### S4 Code and Data Availability

All analysis code, the exact random seeds for every cross-validation split and bootstrap resample, and the scripts that generated every table and figure in this Supplementary Information are available in the public repository accompanying this manuscript (<https://github.com/Alon-Gorenshtein/LLM-BCI-Fusion>). Selections, neural posteriors, and language-model prior posteriors are read-only inputs materialized by the companion attribution study; they will be deposited in a public Zenodo archive with a citable DOI upon acceptance and are not otherwise redistributed here. Access to the underlying BigP3BCI archive is open and requires no data use agreement.
